# *HFE* genotypes, haemochromatosis diagnosis and clinical outcomes to age 80: a prospective cohort study in UK Biobank

**DOI:** 10.1101/2023.11.17.23298637

**Authors:** Mitchell R Lucas, Janice L Atkins, Luke C Pilling, Jeremy Shearman, David Melzer

**Affiliations:** Epidemiology and Public Health Group, Department of Clinical and Biomedical Sciences, Faculty of Health and Life Sciences, University of Exeter, UK; Department of Gastroenterology, South Warwickshire University NHS Foundation Trust, UK

**Keywords:** Haemochromatosis, *HFE* p.C282Y / p.H63D genotypes, iron overload, morbidity, mortality, UK Biobank

## Abstract

**Objectives:** *HFE* haemochromatosis genetic variants have an uncertain clinical penetrance, especially to older ages and in undiagnosed groups. We estimated p.C282Y and p.H63D variant cumulative incidence of multiple clinical outcomes in a large community cohort.

**Design:** Prospective cohort study.

**Setting:** 22 assessment centres across England, Scotland, and Wales in the UK Biobank (2006-2010).

**Participants:** 451,270 participants genetically similar to the 1000-Genomes European reference population, with a mean 13.3-year follow-up through hospital inpatient, cancer registries and death certificate data.

**Main outcome measures:** Cox proportional hazard ratios of incident clinical outcomes and mortality in those with *HFE* p.C282Y-p.H63D mutations compared to those with no variants, stratified by sex and adjusted for age, assessment centre and genetic stratification. Cumulative incidences were estimated from age 40 to 80 years.

**Results:** 12.1% of p.C282Y+/+ males had baseline (mean age 57) haemochromatosis diagnoses, with age 80 cumulative incidence of 56.4%. 33.1% died vs. 25.4% without *HFE* variants (Hazard Ratio [HR] 1.29, 95% CI: 1.12-1.48, p=4.7*10^-4^); 27.9% vs 17.1% had joint replacements, 20.3% vs 8.3% had liver disease, and there was excess delirium, dementia, and Parkinson’s disease, but not depression. Associations, including excess mortality, were similar in the group undiagnosed with haemochromatosis. 3.4% of p.C282Y+/+ females had baseline haemochromatosis diagnoses, with cumulative age 80 incidence of 40.5%. There was excess incident liver disease (8.9% vs 6.8%; HR 1.62, 95% CI: 1.27-2.05, p=7.8*10^-5^), joint replacements and delirium, with similar results in the undiagnosed. p.C282Y/p.H63D and p.H63D+/+ men or women had no statistically significant excess fatigue or depression at baseline and no excess incident outcomes.

**Conclusions:** Male and female p.C282Y homozygotes experienced greater excess morbidity than previously documented, including those undiagnosed with haemochromatosis in the community. As haemochromatosis diagnosis rates were low at baseline despite treatment being considered effective, trials of screening to identify people with p.C282Y homozygosity early appear justified.

**Strengths and limitations of this study:**

- We analyzed largescale data on community volunteers from the UK Biobank, one of the world’s largest *HFE* genotyped cohorts.
- We have analyzed incident disease outcomes during an extended follow-up period of mean 13.3 years.
- We have provided the first clinical outcome data to age 80 years in those with haemochromatosis genotypes, including those undiagnosed with haemochromatosis at baseline, expanding the life-course evidence on *HFE* penetrance.
- UK Biobank participants were somewhat healthier than the general population, but *HFE* allele frequencies were similar to previous UK studies.
- Incident outcomes were from hospital inpatient and cancer registry follow-up, so did not rely on potentially biased patient self-reporting, but community diagnosed conditions may be underestimated.

## Introduction

*HFE* haemochromatosis is defined by iron overload[1] [2] due to gene variants, which dysregulate intestinal iron absorption. The p.C282Y+/+ (homozygous) group have markedly raised iron measures: e.g. median transferrin saturations were over 80% and near 60% in p.C282Y+/+ males and females in HEIRS study, but below 45% in compound heterozygotes (C282Y+/H36D+), and progressively lower across p.H63D+/+ (homozygote), p.C282Y+/- and p.H63D+/- carriers[3]. Women with each genotype have lower mean iron measures than men[4].

Clinical presentation of haemochromatosis is usually with fatigue, joint pain, raised iron measures or from family screening, or less commonly from direct-to-consumer genotyping. Symptoms usually present after age 40 years[5]. In severely affected p.C282Y+/+ patients (>90% of typical cases[6]), liver iron deposition can lead to liver fibrosis, cirrhosis and cancer[7], especially in the presence of other causes of liver disease. Arthropathy[8], diabetes[9], endocrine dysregulation[10], heart arrhythmias and cardiomyopathies[11], and pneumonia[9] have also been reported.

While clinical cohorts frequently have iron overload complications, disease penetrance in population genotyped groups is uncertain, especially at older ages and those not diagnosed with haemochromatosis[3] [12] [13]. Beutler et al found negligible haemochromatosis symptoms in 152 p.C282Y homozygotes from California health appraisal clinics (excluding diagnosed patients)[12]. The HEIRS Study reported excess liver disease in 299 p.C282Y+/+ males[14]. The Melbourne Collaborative Cohort Study[15] reported that 28.4% (95% Confidence Interval [95% CI], 18.8% to 40.2%) of p.C282Y+/+ males (n=95, mean age 65 at follow-up) had ‘documented iron-overload-related disease’, with 1.2% of p.C282Y+/+ females affected. Similarly, although excess mortality occurred in clinical patients (especially with liver disease)[16][17], no excess mortality was reported in community-identified p.C282Y+/+ males or other *HFE* genotype groups[15] [18]. Using UK Biobank, we previously examined data on European ancestry community participants with mean 7-year follow-up, finding the 1,294 male p.C282Y homozygotes had increased odds of liver disease and osteoarthritis compared to those without p.C282Y or p.H63D variants[9]. A UK Biobank 8.9-year follow-up quantified excess hepatic malignancies in male p.C282Y homozygotes (HR 10.5; 95% CI: 6.6-16.7; p<0.001 versus no *HFE* variant) and excess all-cause mortality (n=88 deaths; HR 1.2; 95% CI: 1.0-1.5; p=0.046)[7].

For p.C282Y/H63D compound heterozygotes, the Melbourne Collaborative Cohort Study[15] found only one male (of 242 studied) with documented iron overload-related disease, although alcohol was also a factor. Similarly, a study of community identified participants with p.H63D variants from the Busselton study (Australia) found none with clinically significant iron overloading[19].

Given the accumulating evidence of significant clinical penetrance with p.C282Y homozygosity (only) and the reported effectiveness of treatment (predominantly venesection), there is renewed interest in screening for those at high risk[1] [20] [21]. *HFE* p.C282Y homozygosity is recommended for reporting to patients when found incidentally[22].

The UK Biobank community cohort includes baseline questionnaires, plus hospital inpatient diagnoses during the mean 13.3-year follow-up. The cohort therefore provides a potential model for community based genetic screening. Participant consent did not allow genotype feedback (see Methods), so outcomes reflect normal clinical care. Here we aimed to estimate risks and cumulative outcomes to age 80 years by genotype and sex for relevant clinical outcomes, including (for the first time) analyses of those undiagnosed with haemochromatosis at baseline.

## Methods

### Study population

UK Biobank includes community volunteers aged 39 to 73 years at baseline assessments across England, Scotland and Wales, from 2006 to 2010. Participants were somewhat healthier than the general population[23], but *HFE* allele frequencies were similar to previous UK studies[7]. Data cover 451,270 participants, genetically similar to the 1000 Genomes project European reference population[24], with *HFE* p.C282Y (rs1800562) and *HFE* p.H63D (rs1799945) genotypes from whole exome sequencing (Whole Exome Sequence methods were by Regeneron[25]). Participants gave written informed consent and were informed of relevant health related findings at baseline, but consent excluded individual notification of subsequent findings including genotypes. North West Multi-Centre Research Ethics Committee (Research Ethics Committee reference 11/NW/0382) approved UK Biobank. All research was conducted in accordance with both the Declarations of Helsinki and Istanbul.

### Baseline variables and incident health outcomes

Baseline questionnaires covered doctor-diagnosed conditions including haemochromatosis. Symptom questions included: “Over the past two weeks, how often have you felt tired or had little energy?”, and responses were coded as ‘fatigue’ combining “more than half the days” and “nearly every day”. Studied diagnoses were from *a priori* knowledge (see Supplementary eTable 1 for ascertainment codes for 34 outcomes). England, Wales, and Scotland hospital records were available from April 1996 to October 2022. Prevalent diagnoses were from baseline self-report plus hospital inpatient data from 1996 to baseline. Incident diagnoses and surgical procedures were from hospital inpatient data (baseline to October 2022) plus cancer registries to December 2020 for England and Wales and November 2021 for Scotland. National death records were available to November 2022. Disease ascertainment used International Classification of Diseases 10^th^ revision (ICD-10) codes. Surgical procedures were from OPCS Classification of Interventions and Procedures version 4 (OPCS-4). Having ‘any joint replacement surgery’ included hip, knee, ankle, or shoulder replacement surgery. The ‘any brain outcome’ included delirium, dementia, or Parkinson’s disease.

### Statistical analysis

Cox proportional hazards regression estimated genotype associations with incident outcomes. Models were stratified by sex and adjusted for age, assessment centre and 10 genetic principal components (accounting for population substructure). Main outcome proportional hazards assumptions (tested using ‘estat phtest’) were met in models adjusted for 5-year age bands. Given the extensive prior evidence for risks in p.C282Y+/+ groups, multiple testing corrections are discussed for lower risk genotypes, using Bonferroni correction of p<0.001 (0.05 / 34 disease outcomes). Kaplan-Meier survivor functions estimated the probabilities of cumulative incidence for associated outcomes from age 40 to 80 years within 5-year bands, by *HFE* genotype and by sex. We applied observed incidence rates in each age group to a notional cohort, estimating hypothetical cumulative incident case numbers from age 40 to 80 years. Sensitivity analysis repeated main analyses excluding participants with a haemochromatosis diagnosis at baseline. All analyses were performed in Stata 17.0.

### Patient and public involvement

Patients and participants were and are extensively involved in the UK Biobank study itself. We used anonymised data that were already collected and therefore no patients were involved in developing the research question or the outcomes tested. UK Biobank notified participants of relevant health related findings in the baseline assessment, but there is no individual notification of subsequent findings, including genotypes.

## Results

UK Biobank baseline characteristics were previously reported[9] [26] (Table 1, eTables 2-3): 451,270 participants genetically similar to the European 1000 Genomes reference population were followed for a mean 13.3 years. There were 1,298 p.C282Y+/+ (homozygotes), 4,959 p.C282Y/p.H63D compound heterozygote, and 4,673 p.H63D+/+ males: for females 1,604, 5,760 and 5,580 respectively. In the male p.C282Y+/+ group, 12.1% had haemochromatosis diagnoses at baseline, with 6/1,000 diagnosed in p.C282Y/H63D and 2 per 1,000 in the p.H63D+/+ group: the respective rates in females were 3.4%, 2 per 1,000 and 4 per 10,000.

**Table 1.**
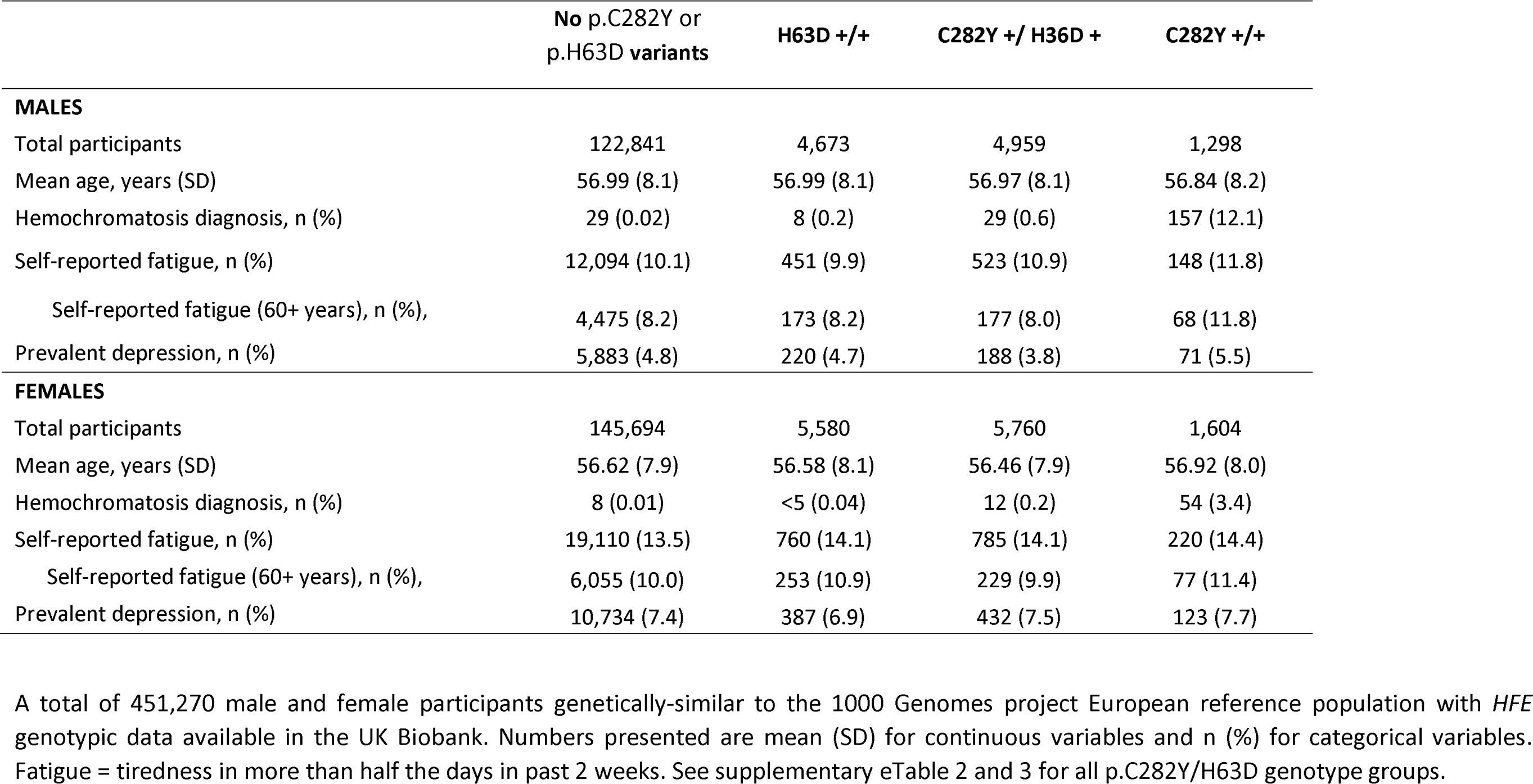
Baseline characteristics of male and female UK Biobank participants by selected p.C282Y/H63D genotypes.

p.C282Y+/+ males aged 60 plus reported baseline excess fatigue (11.8% vs 8.2%; Odds Ratio (OR): 1.43, 95% CI: 1.11-1.85, p=0.01), but there was no statistically significant excess fatigue with other genotypes, except a marginal association in p.H63D+/- males (OR: 1.06, CI 1.00-1.12, p=0.05) which became non-significant after multiple testing correction. There were no differences in depression prevalence at baseline (Table 1, eTables 2-3).

### Males with p.C282Y homozygote genotypes: excess mortality and morbidity versus without HFE variants

Figure 1 shows the Hazard ratios (HRs) for studied incident outcomes by sex and genotype, with cumulative incidence (with confidence intervals) to age 80 years presented in Figure 2 for those outcomes which had significant HRs. p.C282Y+/+ males had increased rates of mortality versus those without p.C282Y or p.H63D variants (HR 1.29, 95% CI: 1.12-1.48, p=4.7*10^-4^; Figure 1; eTables 4 and 5). Cumulative incidence of death was 33.1% (95% CI: 28.9% to 37.8%) versus 25.4% (Figure 2 and 3a). Excess mortality in those undiagnosed with haemochromatosis at UK Biobank baseline (eTables 6 and 7) was similar (HR=1.22 95% CI: 1.05-1.43, p=1.0*10^-2^) with a cumulative death rate of 32.5% (95% CI: 27.9 to 37.6%) (eFigure 1a).

**Figure 1.**
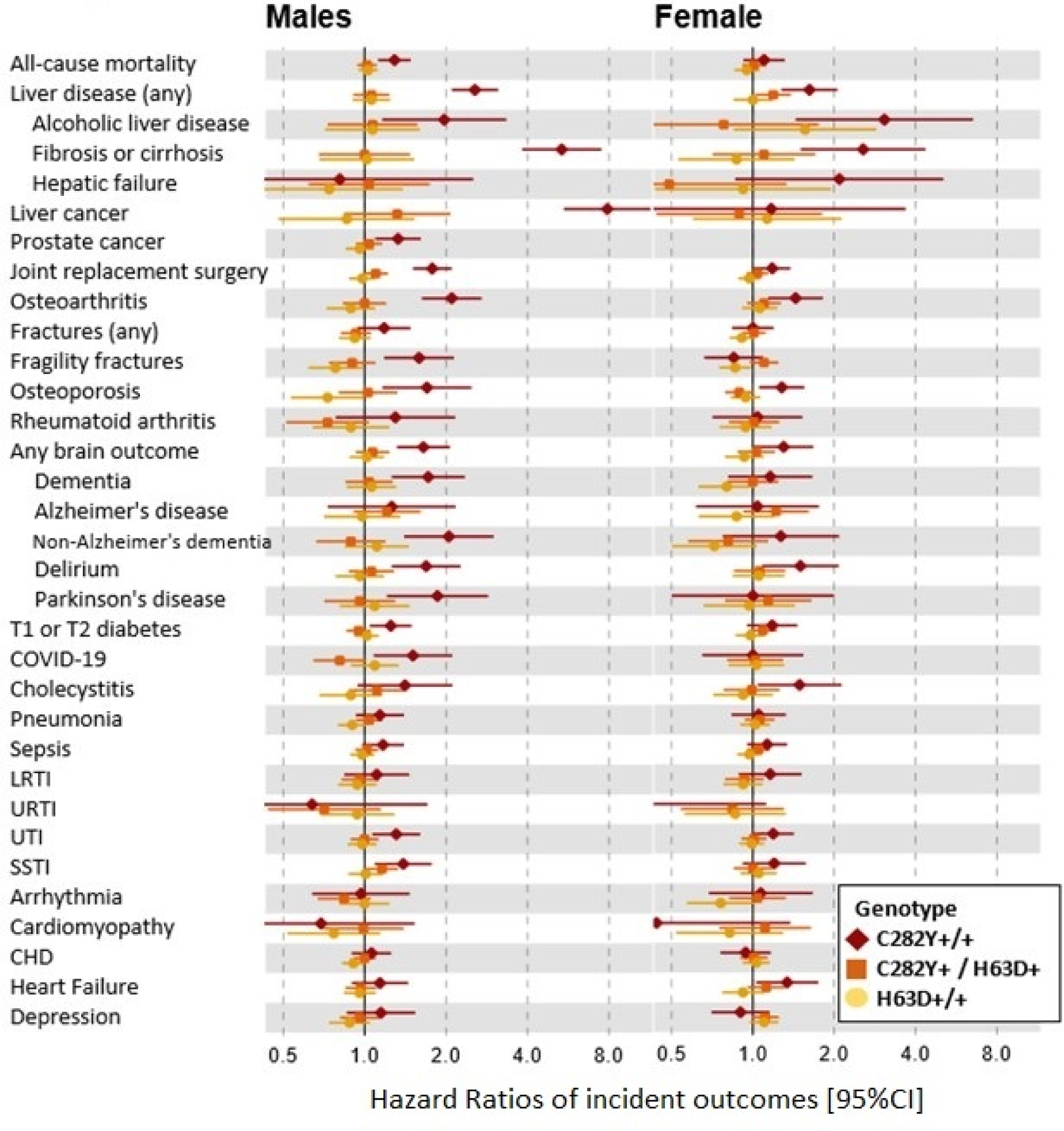
Hazard ratios of incident disease outcomes (95% CI) in selected p.C282Y/H63D genotypes compared to those with no mutations. Hazard ratios compared to those with neither *HFE* mutation. Cox proportional hazards regression models adjusted for age, assessment centre, and genetic principal components 1–10. Abbreviations: UTI, urinary tract infection; SSTI, skin and soft tissue infection; CI, confidence interval. Joint replacement surgery includes a diagnosis of hip, knee, ankle, or shoulder replacement. Any brain outcome included a diagnosis of delirium, dementia, or Parkinson’s disease. See supplementary eTables 4, 5, 9 and 10 for incident numbers and HRs for all p.C282Y/H63D genotype groups.

**Figure 2.**
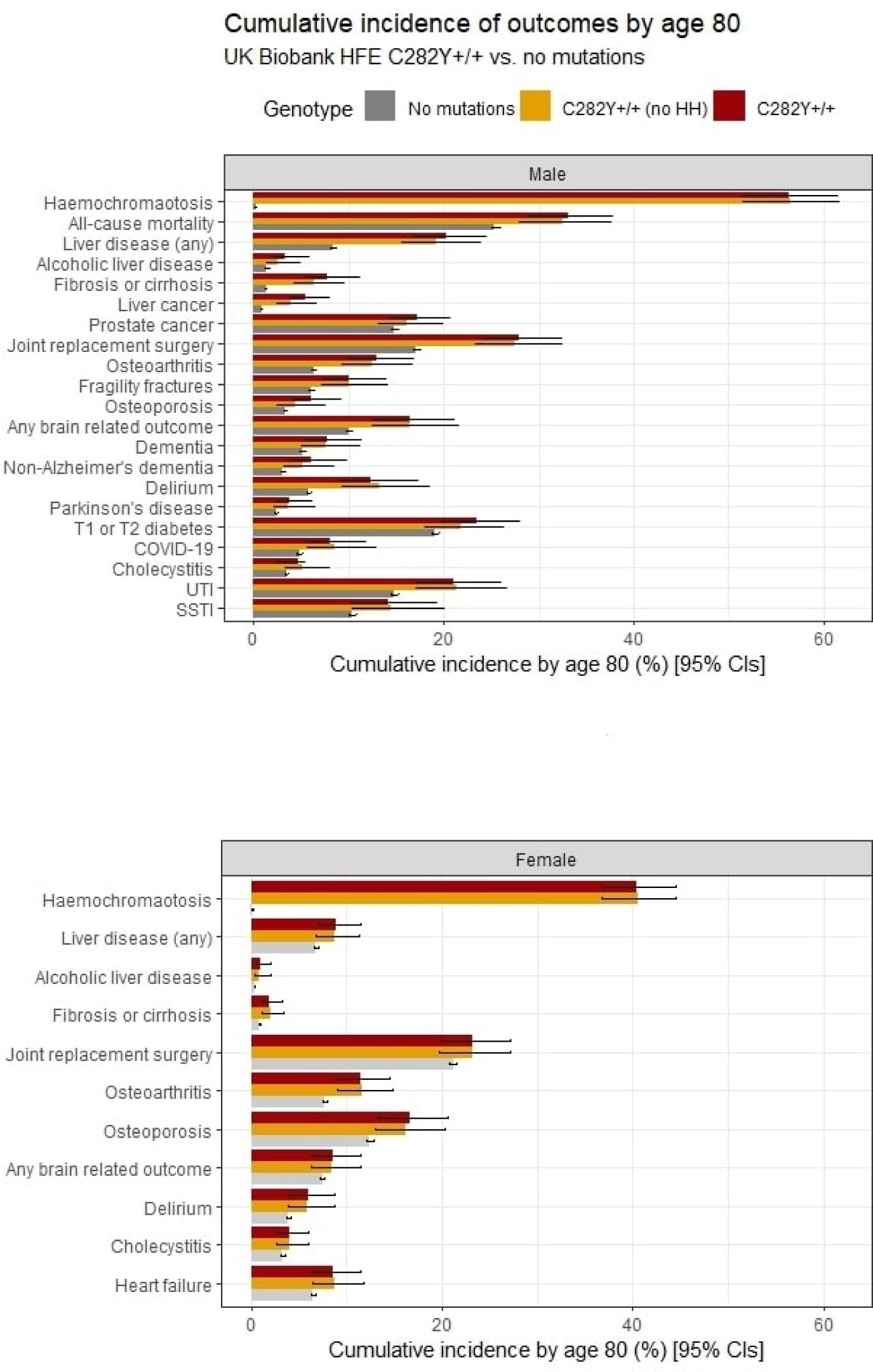
Cumulative incidence of outcomes from ages 40-80 years by *HFE* genotypes (no p.C282Y or p.H63D variants vs p.C282Y homozygotes and p.C282Y homozygotes undiagnosed with hemochromatosis at baseline). Cumulative incidence (estimated % diagnosed by age 80 years; 95% CIs) for significant outcomes (p<0.05) from Cox proportional hazards regression models (Figure 1; eTables 5, 7, 10 and 12). Abbreviations: UTI, urinary tract infection; SSTI, skin and soft tissue infection; CI, confidence interval. Joint replacement surgery includes a diagnosis of hip, knee, ankle, or shoulder replacement. Any brain outcome included a diagnosis of delirium, dementia, or Parkinson’s disease.

Haemochromatosis diagnosis cumulative incidence at age 80 in p.C282Y+/+ males was 56.4% (Figure 2, eTable 8). Cumulative incidence of ‘any liver disease’ was 20.3% vs 8.3% without variants (HR 2.56, 95% CI: 2.10-3.12, p=8.70*10^-21^) and 7.7% developed liver fibrosis or cirrhosis vs 1.3%. There was a raised HR for alcoholic liver disease (Figure 1) with cumulative incidence 3.3% of p.C282Y+/+ males vs 1.4%. Liver cancers cumulative incidence was 5.5% (95% CI: 3.8% to 8.0%, vs 0.8% without variants). p.C282Y+/+ males also had raised cumulative incidence of prostate cancer: 17.2% vs 14.8%. The cumulative incidence graphs show excess liver disease clearly apparent by age 55, but the excess mortality becoming significant at older ages (Figure 3a and 3b). Notably, 81.5% of deaths, 66.3% of ‘any liver diseases’, and 71.8% of joint replacements in p.C282Y+/+ males occurred after age 65.

**Figure 3.**
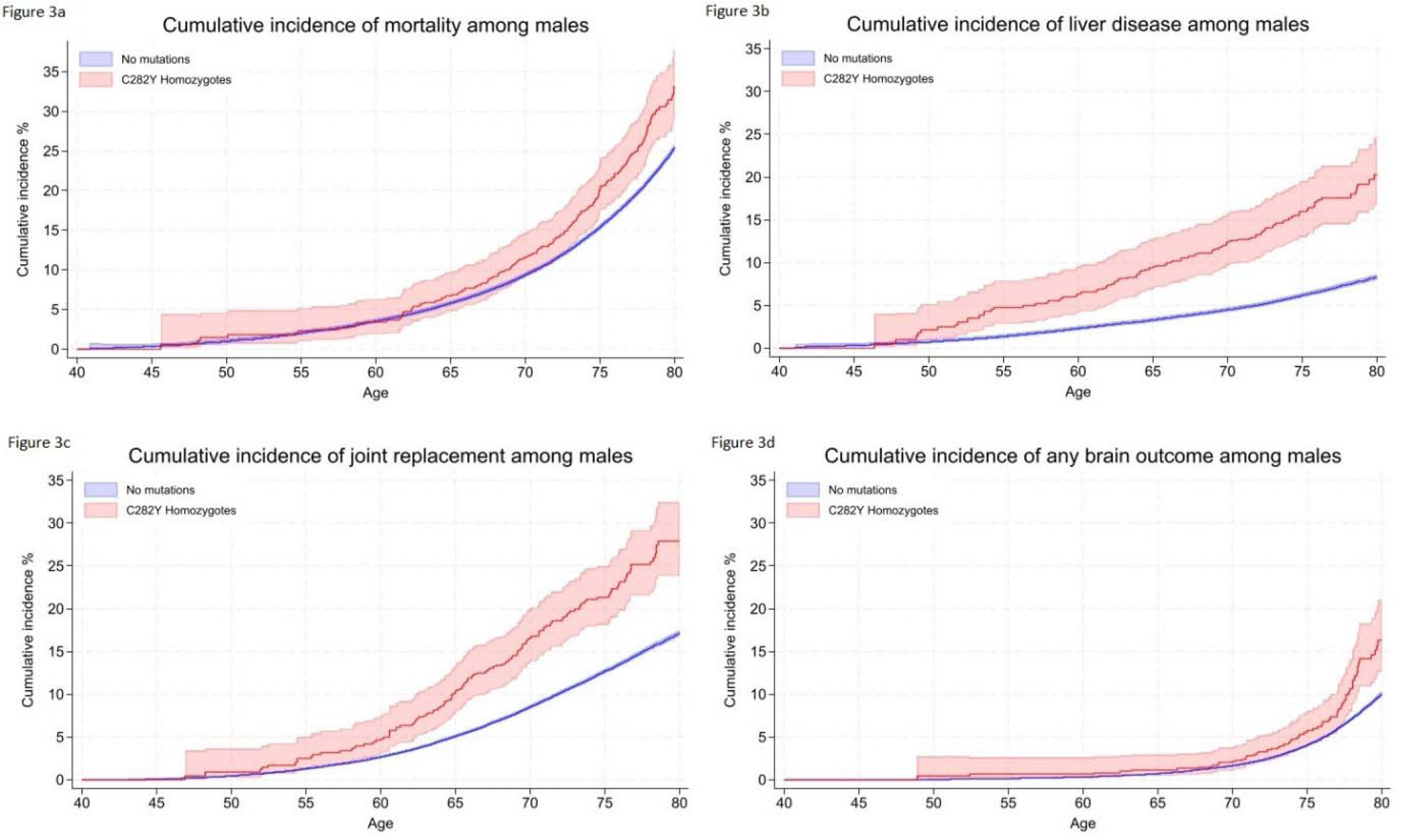
Kaplan-Meier curves for the cumulative incidence of (a) mortality, (b) liver disease, (c) any joint replacement, and (d) any brain outcome, in male *HFE* p.C282Y homozygotes compared to those with no mutations. Cumulative incidence (estimated % diagnosed by age 80 years; 95% CIs). Joint replacement surgery includes a diagnosis of hip, knee, ankle, or shoulder replacement. Any brain outcome included a diagnosis of delirium, dementia, or Parkinson’s disease.

Joint replacement cumulative incidence in p.C282Y+/+ males was 27.9% vs 17.1% without variants (Figure 3c): this remained after excluding those with fractures within 5 days before surgery (n=53; HR 1.74, 95% CI: 1.45-2.09; p=1.6*10^-9^). p.C282Y+/+ males also had excess osteoarthritis, fragility fractures, and osteoporosis.

Brain outcome (dementia, delirium, or Parkinson’s disease) cumulative incidence in p.C282Y+/+ males was 16.3% versus 10.0% without variants (HR 1.65 95% CI: 1.31-2.06, p=1.70*10^-5^) (Figure 3d); for delirium 12.4% vs. 5.8% (HR 1.69, 95% CI: 1.26-2.27; p=4.80*10^-4^); non-Alzheimer’s dementia 6.0% vs 3.0% (HR 2.05, 95% CI: 1.40-3.00; p=2.40*10^-4^), and Parkinson’s disease (HR 1.86, 95% CI: 1.21-2.87 p=0.005).

Diabetes (type 1 or 2) cumulative incidence was 23.5% in p.C282Y+/+ males (vs. 19.1%). p.C282Y+/+ males had excess hospital diagnosed COVID-19 (8% vs 4.8%, HR 1.51, 95% CI: 1.08-2.11; p=0.02, missing significance in earlier follow-up data[27]), urinary tract infections (UTI) and skin and soft tissue infections (SSTI). Associations were not significant for cardiac outcomes (arrhythmia, cardiomyopathy, CHD, heart failure) or depression.

Estimates of excess morbidity in the male p.C282Y+/+ group without haemochromatosis diagnoses in the community at UK Biobank baseline (n=1,141) were mostly similar to those in the overall sample (Figures 2 and eFigure 1a-d, eTables 6 and 7). Notably, however, point HR estimates for liver fibrosis or cirrhosis, and liver cancers risk appeared marginally lower than in the whole group, but with wide confidence intervals including the whole study estimates HR point estimates (e.g. for liver fibrosis and cirrhosis HR 4.52, 95% CI: 3.07-6.66, p=2.10*10^-14^ without diagnosis, versus HR 5.36 95% CI: 3.83-7.52; p=2.00*10^-22^ in the whole p.C282Y+/+ group). In addition, alcoholic liver disease and osteoporosis lost statistical significance, but cholecystitis became nominally statistically significant (HR 1.54, 95% CI: 1.02-2.32, p=0.04).

### Females with p.C282Y homozygote genotypes: excess morbidity versus without HFE variants

Diagnosed haemochromatosis cumulative incidence in p.C282Y+/+ females was 40.5% by age 80. There was no excess mortality, but this group experienced excess ‘any liver disease’ (8.9% vs 6.75%; HR 1.62, 95% CI: 1.27-2.05; p=7.80*10^-5^) (Figure 4a), liver fibrosis or cirrhosis (1.9% vs 0.8%; HR 2.56, 95% CI: 1.50-4.36; p=0.001), alcoholic liver disease (1.0% vs 0.3%; HR 3.07, 95% CI: 1.44-6.54; p=0.004), plus cholecystitis, versus women without *HFE* variants (Figures 1 and 2; eTables 8, 9 and 10).

**Figure 4.**
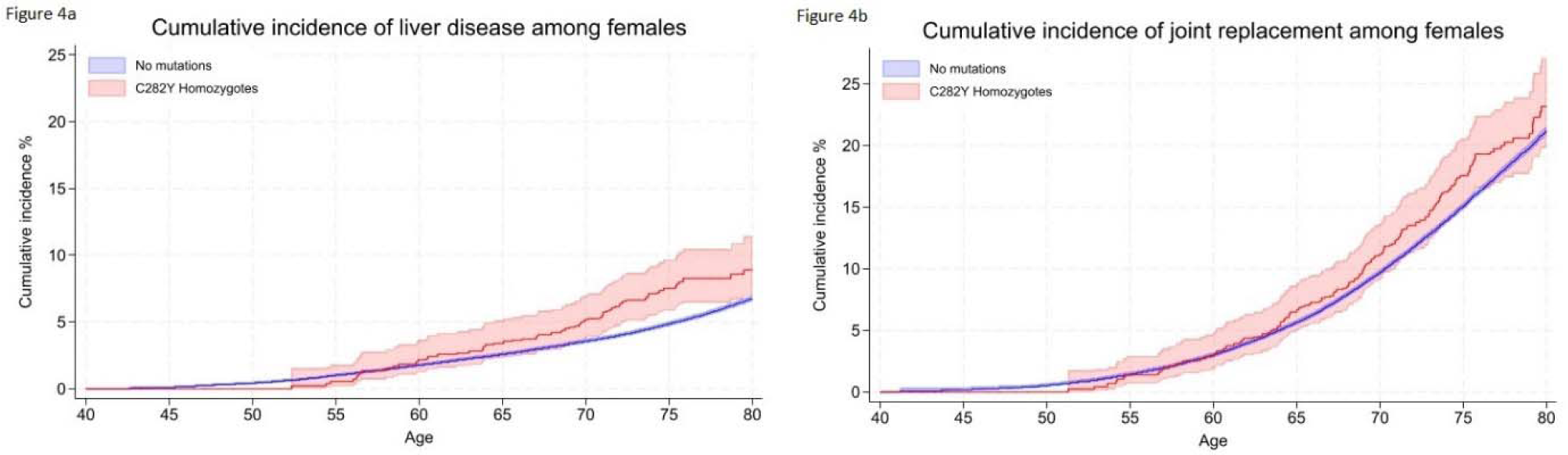
Kaplan-Meier curves for the cumulative incidence of (a) liver disease, and (b) any joint replacement, in female *HFE* p.C282Y homozygotes compared to those with no mutations. Cumulative incidence (estimated % diagnosed by age 80 years; 95% CIs). Joint replacement surgery includes a diagnosis of hip, knee, ankle, or shoulder replacement.

Joint replacement surgery was more common in female p.C282Y homozygotes (23.2% vs 21.1%) (Figure 4b), as were osteoarthritis and osteoporosis. Cumulative incidence of brain outcomes was raised (8.6% vs 7.4%; HR 1.30, 95% CI: 1.01-1.68; p=0.04), including delirium (5.9% vs 3.9%; HR 1.50. 95% CI: 1.08-2.08; p=0.02).

There was a nominally significant excess of heart failure (HR 1.34, 95% CI:1.03-1.75, p=0.03) but no associations with other studied cardiac outcomes. There were no associations with liver cancer, fragility fractures, diabetes, dementia, Parkinson’s disease, or infections (SSTIs, UTIs).

Estimates of excess morbidity in the female p.C282Y+/+ group without haemochromatosis diagnoses in the community at UK Biobank baseline (n=1,550) were similar to those in the overall sample (Figures 2 and eFigure 2a-b, eTables 11 and 12).

### Male and female p.C282Y/p.H63D compound heterozygotes versus without HFE variants

Haemochromatosis diagnosis cumulative incidence in p.C282Y/H63D males was 5.4% by age 80. However, there was no statistically significant increase in mortality, incidence of ‘any liver disease’ or liver cancers, joint replacements, or diabetes (Figure 1). p.C282Y/H63D males were modestly more likely to develop SSTIs (HR 1.16, 95% CI: 1.01-1.33, p=0.03) but this lost statistical significance after multiple testing correction.

Haemochromatosis cumulative incidence in p.C282Y/H63D females was 2.7% by age 80 but there were similarly no excess mortality, musculoskeletal diagnoses, diabetes, or brain outcomes (Figure 1). However, there was a modest association with ‘any liver disease’ (HR 1.19, 95% CI: 1.02-1.38, p=0.03), but associations with specific liver diagnoses were non-significant, including for liver fibrosis and cirrhosis, the most common form of liver disease with iron overload (Figures 1-2; eTables 4, 5, 8, 9, 10).

### Males and females with p.H63D homozygote genotypes vs. no HFE variants

Diagnosed haemochromatosis cumulative incidence in p.H63D/H63D males was 1.9% by age 80, but with no excess mortality, liver disease diagnoses, liver cancers, joint replacement surgery, diabetes, or depression. There were protective associations between p.H63D homozygosity and fragility fractures (HR 0.78 95% CI: 0.62-0.98 p=0.03) and osteoporosis (HR 0.73 95% CI: 0.53-0.99, p=0.04) (eTables 4, 5 and 8).

Diagnosed haemochromatosis cumulative incidence p.H63D/H63D females was 0.6% by age 80. There was no excess mortality, liver disease or liver cancers, joint replacement surgery, diabetes, or depression. However, females also had a nominally significant (uncorrected p<0.05) protective association with fragility fractures (HR 0.87 95% CI: 0.76-1.00 p=0.05), echoing the similar protective associations in p.H63D/H63D males (eTables 8, 9 and 10).

### Associations for p.C282Y+/- and p.H63D+/- (heterozygote carrier) males and females

There were a small number of nominally significant protective and risk associations in carrier groups (eTables 4 and 5), in p.C282Y+/- males including for excess delirium (HR 1.13 95% CI: 1.04-1.24 p=0.006) and skin and soft tissue infections (HR 1.14 95% CI: 1.06-1.22 p=1.80*10^-4^) and for p.H63D+/- males a protective association with fragility fractures (HR 0.92 95% CI: 0.86-0.99 p=0.03). For p.C282Y+/- females there were no associations, but for p.H63D+/- females a nominally significant excess of liver cancers (HR 1.26 95% CI: 1.01-1.58 p=0.04) was present, plus a protective association for osteoporosis (eTables 9 and 10). Only the skin and soft tissue infection association was significant after multiple testing correction.

## Discussion

Clinical penetrance by *HFE* genotypes in community samples has been uncertain, especially for those undiagnosed for haemochromatosis, the potential beneficiaries of population screening. Our results show greater excess morbidity and mortality in p.C282Y+/+ groups than previously documented, much occurring at older ages. p.C282Y+/+ males had marked excess mortality (33.1% dead by age 80 vs. 25.4% without p.C282Y or p.H63D variants), plus substantial excess joint replacements, liver disease, brain outcomes and some infections. Estimates of excess morbidity in p.C282Y+/+ groups undiagnosed for haemochromatosis were similar (eTables 7 and 12): perhaps most crucially, the p.C282Y homozygote male undiagnosed group had similar excess mortality, with an estimated cumulative death rate to age 80 of 32.5% (95% CI: 27.9 to 37.6%) versus 25.4% in those without *HFE* variants (eFigure 1a and eTable 7). Of p.C282Y+/+ females (n=1,604), diagnosed haemochromatosis cumulative incidence by 80 was 40.5%, with excess ‘any liver disease’, joint replacements and delirium present, suggesting more clinical penetrance than previously documented. In p.C282Y/p.H63D and H63D homozygote groups and carriers, we found no excess fatigue or depression at baseline and no excess mortality or major outcomes during follow-up.

### Comparison to previous fatigue and depression studies

Baseline characteristics in UK Biobank have been reported before,[26], except for fatigue and depression. Fatigue and depression in haemochromatosis is important to patients and their carers, but the causal role of iron overload in these symptoms is unclear. p.C282Y+/+ males aged 60 plus reported excess fatigue (11.8% versus 8.2% without *HFE* variants) at baseline, but no excess depression at baseline or follow-up. Rates of both conditions were similar in the other studied genotype groups to those without *HFE* variants (except in p.H63D+/- males, with very modest excess fatigue, non-significant with multiple statistical testing correction). A blinded randomized trial of erythrocytapheresis[28] in p.C282Y+/+ groups with moderately raised ferritin levels (300 and 1000 µg/L) did find reduced fatigue scores with transferrin saturations falling from mean 63.5% to 45.4% in the treatment group, although the mean fatigue score was below diagnosis level, making clinical interpretation unclear. Also, recent observational studies have cast doubt on relationships between fatigue, quality of life measures and iron parameters in haemochromatosis patients[29] [30], and fatigue might also occur with venesection. A UK biobank analysis found tiredness genetically linked to factors including increased adiposity, blood lipids and inflammatory markers[31].

### Comparison to previous mortality-and morbidity studies

Rates of haemochromatosis diagnosis at baseline (mean age 57) were low even in p.C282Y+/+ groups (12.1% in males, 3.4% in females), but cumulative estimates to age 80 indicated much higher diagnosis rates at older ages. These baseline rates are similar to cumulative rates from the US hospitals genetics collaboration eMERGE study[32] for p.C282Y+/+ group at age 60. The eMERGE study[32] figures also show the majority of haemochromatosis diagnoses occurring later in life, reaching nearly 50% in males and 25% in females after age 80.

Earlier reports suggested no excess mortality in community-identified p.C282Y+/+[15] [18], despite higher death rates in clinical cohorts with liver disease[33], [15]. In the UK Biobank much larger sample, we found clearly increased all-cause mortality in p.C282Y+/+ males only (194 p.C282Y+/+ deaths, HR 1.29, 95% CI: 1.12 to 1.48, p=4.70*10^-4^). A Swedish sample of 2,273 p.C282Y homozygotes found similar increased mortality (HR 1.30, 95% CI: 95%: 1.12-1.50, versus males with no mutations), with no excess in female homozygotes (HR 0.98, 95% CI: 0.82-1.18)[34]. These studies found no excess mortality in p.C282Y/p.H63D or p.H63D/p.H63D groups.

Estimates of disease penetrance in community genotyped groups differ widely [3][12] [13], with the highest estimate from The Melbourne Collaborative Cohort Study[15], of 28.4% (95% CI: 18.8% to 40.2%) of p.C282Y+/+ males (n=95, mean age 65 at follow-up) having ‘documented iron-overload-related disease’, and 1.2% of female p.C282Y homozygotes affected. In UK Biobank we found that diagnosed haemochromatosis cumulative incidence was 36.1% in p.C282Y+/+ males and 21.2% of p.C282Y+/+ females by age 65 (eTable 8), suggesting a larger burden of iron overload disease especially in p.C282Y+/+ women. Liver cancers occurred in 5.5% (95% CI: 3.8% to 8.0%) of p.C282Y+/+ males by age 80, a slightly lower estimate than at age 75[7] (7.2% (95% CI: 3.9%-13.1%), but within the earlier confidence intervals. The estimate remains comparable to a meta-analysis[13] estimated lifetime incidence of severe liver disease (cirrhosis or hepatocellular carcinoma) of 9% (95% CI: 2.6%-15.3%) in untreated male *HFE* p.C282Y+/+ homozygotes.

Several studies have reported excess arthritis in p.C282Y homozygote men and to a lesser extent, in female homozygotes[9]. Our results extend a previous 11.5-year follow-up[35], now showing the full extent of later-life joint replacements in male p.C282Y homozygotes and providing a robust measure for severe joint damage in haemochromatosis.

Excess diabetes has previously been reported in p.C282Y homozygous men (9), but other studies have suggested that *HFE* mutations do not have important pathophysiological consequences in patients with type 2 diabetes[36]. We found a modest diabetes excess in p.C282Y+/+ individuals (Figures 1 and 2). Overall, as cumulative diabetes incidence was 23.5% in p.C282Y+/+ males vs. 19.1% without *HFE* variants, non-iron factors appear to now contribute the majority of diabetes even in p.C282Y+/+ males. Although cardiac complications are often noted haemochromatosis reviews[11][37], there is little evidence linking community-identified p.C282Y homozygosity to these outcomes[38] [39]: our finding of excess of heart failure in p.C282Y homozygote women only (HR 1.34 95% CI: 1.03-1.75, p=0.03) needs further study.

Our previous UK Biobank report on brain outcomes (mean 10.5 years follow-up) found p.C282Y+/+ males had excess dementia diagnoses and iron deposition in key brain areas on MRI[40]. The current analysis also found higher incidence especially of non-Alzheimer’s dementia in p.C282Y+/+ males plus more delirium. Loughnan et al[41] reported that p.C282Y+/+ males had more Parkinson’s disease (OR 1.83; 95% CI: 1.19-2.80; p=0.006) in cross-sectional analyses of UK Biobank: our incident analyses provide a similar estimate for Parkinson’s disease in p.C282Y+/+ males (HR 1.86, 95% CI: 1.21-2.87 p=0.005).

In the HEIRS study[3] community genotyped p.C282Y/p.H63D (compound heterozygote) males and females had substantially lower transferrin saturation levels compared to p.C282Y+/+ males or females. In line with this, cumulative incidence of haemochromatosis diagnosed by age 80 was relatively low (men 5.4%, women 2.7%), and no excess mortality, liver fibrosis and cirrhosis, joint replacements, diabetes, or depression in these groups. We did find that p.C282Y/p.H63D females had nominally significant small excess of any liver disease (HR 1.19, 95% CI: 1.02-1.38; p=0.03) although with no excess liver fibrosis and cirrhosis, no similar association in p.C282Y/p.H63D males, and with a non-significant multiple testing p-value. p.C282Y/p.H63D males had a small excess of skin and soft tissue infections (HR 1.16, 95% CI: 1.01-1.33, uncorrected p=0.03), again multiple testing non-significant.

### Strengths and limitations

We analyzed largescale cohort data on community volunteers, providing the first outcome data to age 80. *HFE* allele frequencies were similar to other UK studies[7] but UK Biobank did recruit somewhat healthier participants than the general population[23], so we have focused on incident outcomes during the mean 13.3 year follow-up: we found no deviations from proportional hazards assumptions in Cox models. Outcomes for both the overall genotype groups and those undiagnosed with haemochromatosis at UK Biobank baseline are presented. Previous studies have suggested that treated p.C282Y homozygous patients do not have an increased mortality rate compared to the general population [42], but numbers with haemochromatosis diagnoses at baseline were small in the current study and so outcomes in this group were not presented. Also, there was no data available on the diverse routes to diagnosis, so modelling treated outcomes would likely be confounded. Further work is needed on the effect so early diagnosis and treatment. Most outcomes were from hospital inpatient care, so community diagnosed conditions may be underestimated, but findings are similar to analyses including primary care records (available to 2017 only)[7] and baseline data collected at interview[9]. Information on the criteria used to establish a diagnosis of fibrosis/cirrhosis in the hospital follow-up data (e.g. non-invasive biomarker panels or liver biopsy) were not available.

### Implications for early diagnosis and screening

We found greater excess morbidity than previously reported in both p.C282Y+/+ males and females, especially at older ages, and in those undiagnosed with haemochromatosis. Despite treatment (predominantly venesection) being considered effective for preventing liver disease progression[1][43][44] rates of haemochromatosis diagnosis were low at baseline: for example, of 31 p.C282Y+/+ males with incident liver cancer, 17 were undiagnosed with haemochromatosis at baseline. Genotyping earlier in life could be a powerful preventive tool to identify p.C282Y+/+ individuals who are clearly at major risk of related excess disease. A recent cross-sectional genotyping study within a US health provider found 72% (144/201) of p.C282Y homozygous patients (mean age 62) had not been diagnosed with haemochromatosis, 36% of whom had iron overload [21]. As cumulative diagnosed haemochromatosis in UK Biobank cohort was estimated 56.4% in p.C282Y+/+ males and 40.5% females by age 80, p.C282Y+/+ genotyping with follow-up iron studies would likely have a high yield. However, the prevention potential following up p.C282Y/p.H63D, H63D+/+ groups will likely be very low, as there was no significant excess fatigue or depression at baseline, and no excess mortality, incident liver fibrosis or cirrhosis, joint replacements, or depression.

## Conclusion

Male and female p.C282Y homozygotes in this community cohort experienced greater excess morbidity than previously documented. Much of this excess liver, musculoskeletal, diabetes, brain and morbidity occurred after age 60. In those not diagnosed with haemochromatosis at study baseline, risks were broadly similar to those in the overall group, notably including excess mortality in p.C282Y+/+ males. Trials of targeted or community genotyping to diagnose haemochromatosis earlier appear justified, especially to identify people with the p.C282Y homozygote variants. The potential for iron related preventive treatment in the p.C282Y/H63D, p.H63D+/+ and other *HFE* genotype groups appears very limited, as these genotype groups were associated with no statistically significant excess morbidity or mortality.

## Data sharing

Data are available on application to the UK Biobank (www.ukbiobank.ac.uk/register-apply).

## Authors contributions

ML performed the analysis, interpreted results, created the figures, and drafted the manuscript. JA contributed to the design and analysis of the study, data interpretation and drafting of the manuscript. LP contributed to the design of the study, data interpretation, creation of figures and contributed to the manuscript. JS provided expert clinical interpretation of the data and contributed to the manuscript. DM oversaw design of the study, data analysis, interpretation of results, and led the writing of the manuscript. All authors approved the final version of the manuscript.

## Data Availability

All data produced in the present study are available upon reasonable request to the authors
All data produced in the present work are contained in the manuscript
This research was conducted using the UK Biobank resource, under application 14631.

## Acknowledgements

This research was conducted using the UK Biobank resource, under application 14631. We thank the UK Biobank participants and coordinators. This work used data provided by patients and collected by the NHS as part of their care and support. Copyright © (2023), NHS England. Re-used with the permission of the NHS England and UK Biobank. All rights reserved. This research also used data assets made available by National Safe Haven as part of the Data and Connectivity National Core Study, led by Health Data Research UK in partnership with the Office for National Statistics and funded by UK Research and Innovation. This study was supported by the National Institute for Health and Care Research Exeter Biomedical Research Centre. The views expressed are those of the authors and not necessarily those of the NIHR or the Department of Health and Social Care.

