## Supplementary Material for "*HFE* genotypes, haemochromatosis diagnosis and clinical outcomes to age 80: a prospective cohort study in UK Biobank"

**eTable 1.** Incident hospital diagnosed outcomes or procedures with associated ICD-10/OPCS codes

| Disease | ICD-10 code |
|---|---|
| Arrhythmia | I49 |
| Cardiomyopathy | I42 |
| CHD | I20; I21; I22; I23; I24; I25 |
| Cholecystitis | K800; K804; K81 |
| COVID-19 | U07.1; U07.2 |
| Delirium | F05 |
| Dementia | F00; F01; F02; F03; G30 |
| Alzheimer's disease | G30 |
| Non-Alzheimer's dementia | F00; F01; F02; F03 |
| Depression | F32; F33; F34.1 |
| Fractures (any) | S02; S12; S22; S32; S42; S52; S62; S72; S82; S92; T02; T08; T10; T12; T14.2 |
| Fragility fractures | S220; S32; S325; S328; S422; S423; S424; S524; S525; S720; S721; S722; S582; S5823; T08 |
| Hemochromatosis | E83.1 |
| Heart failure | I50; J81 |
| Liver disease (any) | K70; K71; K72; K73; K74; K75; K76; K77 |
| Alcoholic liver disease | K70 |
| Fibrosis & Cirrhosis | K74 |
| Hepatic failure | K72 |
| Liver cancer | C22 |
| Lower respiratory tract infection | J20; J21; J22 |
| Osteoarthritis | M15.0; M15.1; M15.2; M15.9; M16.0; M16.1; M17.0; M17.1; M18.0; M18.1; M19.0 |
| Osteoporosis | M80; M81; M811; M812; M813; M814; M815; M816; M818; M819 |
| Parkinson's disease | G20; F02.3 |
| Pneumonia | J13; J14; J15; J16; J17; J18 |
| Prostate cancer | C61 |
| Rheumatoid arthritis | M05; M06 |
| Sepsis | A021; A039; A207; A241; A217; A227; A239; A267; A282; A327; A392; A393; A394; A40; A41; A427; A548; B007; B377; H440; J950; N390; O85; P36; R651; T814; T880 |
| Skin soft tissue infection | L00; L01; L02; L03; L04; L05; L06; L07; L08 |
| Type 1 or Type 2 diabetes | E10; E11 |
| Urinary tract infection | N30; N34; N39 |
| Upper respiratory tract infection | J39; J06; J04 |

| Procedure | OPCS Code |
|---|---|
| Ankle replacement | O32; O320; O321; O322; O323; O324; O325 |
| Hip replacement | W37;W370;W371;W372;W373;W374;W38;W380;W381;W382;W383;W384; W46;W460;W461;W462;W463;W47;W470;W471;W472;W473;W93;W930; W931;W932;W933;W94;W940;W941;W942;W943;O171;O172;O173;W580; W581;W582 |
| Knee replacement | O18*; W40*; W41*; W42* |
| Shoulder replacement | O06; +A11O060; O061; O062; O063; O068; O069; O07; O070; O071; O072; O073; O078; O079; O08; O080; O081; O082; O083; O084; O088; O089; O09; O091; O098; O099; O10; O101; O108; O109 |

ICD-10 = International Classification of Diseases 10th revision codes; OPCS-4 = OPCS Classification of Interventions and Procedures version 4.  Joint replacement surgery variable includes a diagnosis of hip, knee, ankle, or shoulder replacement. Any brain outcome variable includes a diagnosis of dementia, delirium, or Parkinson's disease.

**eTable 2**. Baseline characteristics of male UK Biobank participants by p.C282Y/H63D genotypes

| | No mutations | H63D+/- | H63D+/+ | C282Y+/H63+ | C282Y+/- | C282Y +/+ | Total |
|---|---|---|---|---|---|---|---|
| **Total participants** | 122,841 | 47,983 | 4,673 | 4,959 | 24,636 | 1,298 | 206,390 |
| Mean age, years (SD) | 56.99 (8.1) | 57.02 (8.1) | 56.99 (8.1) | 56.97 (8.1) | 57.02 (8.1) | 56.84 (8.2) | 57.00 (8.1) |
| Hemochromatosis diagnosis, n (%) | 29 (0.02) | 17 (0.04) | 8 (0.2) | 29 (0.6) | 27 (0.1) | 157 (12.1) | 267 (0.1) |
| Self-reported fatigue, n (%) | 12,094 (10.1) | 4,865 (10.4) | 451 (9.9) | 523 (10.9) | 2,476 (10.3) | 148 (11.8) | 20,557 (10.2) |
| Self-reported fatigue (60+ years), n (%), | 4,475 (8.2) | 1,858 (8.6) | 173 (8.2) | 177 (8.0) | 924 (8.4) | 68 (11.8) | 7,675 (8.3) |
| Depression diagnosis, n (%) | 5,883 (4.8) | 2,232 (4.7) | 220 (4.7) | 188 (3.8) | 1,146 (4.7) | 71 (5.5) | 9,740 (4.7) |

A total of 206,390 male participants genetically similar to the 1000 Genomes project European reference population with *HFE* genotypic data available in the UK Biobank. Numbers presented are mean (SD) for continuous variables and n (%) for categorical variables. Fatigue = tiredness in more than half the days in past 2 weeks.

**eTable 3.** Baseline characteristics of female UK Biobank participants by p.C282Y/H63D genotypes

| | No mutations | H63D+/- | H63D+/+ | C282Y+/H63+ | C282Y+/- | C282Y +/+ | Total |
|---|---|---|---|---|---|---|---|
| **Total participants** | 145,694 | 57,021 | 5,580 | 5,760 | 29,221 | 1,604 | 244,880 |
| Mean age, years (SD) | 56.62 (7.9) | 56.58 (7.9) | 56.58 (8.1) | 56.46 (7.9) | 56.49 (8.0) | 56.92 (8.0) | 56.60 (7.9) |
| Hemochromatosis diagnosis, n (%) | 8 (0.01) | 6 (0.01) | 2 (0.04) | 12 (0.2) | 5 (0.02) | 54 (3.4) | 87 (0.04) |
| Self-reported fatigue, n (%) | 19,110 (13.5) | 7,449 (13.5) | 760 (14.1) | 785 (14.1) | 3,802 (13.4) | 220 (14.4) | 32,126 (13.5) |
| Self-reported fatigue (60+ years), n (%), | 6,055 (10.0) | 2,417 (10.2) | 253 (10.9) | 229 (9.9) | 1,157 (9.6) | 77 (11.4) | 10,188 (10.0) |
| Depression diagnosis, n (%) | 10,734 (7.4) | 4,266 (7.5) | 387 (6.9) | 432 (7.5) | 2,271 (7.8) | 123 (7.7) | 18,213 (7.4) |

A total of 244,880 female participants genetically similar to the 1000 Genomes project European reference population with *HFE* genotypic data available in the UK Biobank. Numbers presented are mean (SD) for continuous variables and n (%) for categorical variables. Fatigue = tiredness in more than half the days in past 2 weeks.

**eTable 4**. Incident hospital diagnoses in male UK Biobank participants by p.C282Y/H63D genotypes

| Males | No mutations | H63D +/- | H63D +/+ | C282Y +/ H36D + | C282Y +/- | C282Y +/+ |
|---|---|---|---|---|---|---|
| Hemochromatosis | 85 (0.1) | 51 (0.1) | 29 (0.6) | 95 (1.9) | 42 (0.2) | 288 (25.2) |
| All-cause mortality | 13,976 (11.4) | 5,658 (11.8) | 540 (11.6) | 591 (11.9) | 2,996 (12.2) | 194 (15.0) |
| **Liver** | | | | | | |
| Liver disease (any) | 3,969 (3.3) | 1,621 (3.4) | 160 (3.5) | 171 (3.5) | 803 (3.3) | 102 (8.1) |
| Alcoholic liver disease | 608 (0.5) | 227 (0.5) | 25 (0.5) | 28 (0.6) | 150 (0.6) | 14 (1.1) |
| Fibrosis & Cirrhosis | 618 (0.5) | 252 (0.5) | 24 (0.5) | 26 (0.5) | 142 (0.6) | 36 (2.8) |
| Hepatic failure | 355 (0.3) | 137 (0.3) | 10 (0.2) | 15 (0.3) | 54 (0.2) | 3 (0.2) |
| **Cancer** | | | | | | |
| Liver cancer | 363 (0.3) | 167 (0.4) | 12 (0.3) | 20 (0.4) | 80 (0.3) | 31 (2.4) |
| Prostate cancer | 7,723 (6.4) | 3,025 (6.4) | 283 (6.2) | 319 (6.5) | 1,613 (6.7) | 104 (8.1) |
| **Musculoskeletal** | | | | | | |
| Joint replacement surgery (any) | 8,429 (7.1) | 3,379 (7.3) | 315 (7.0) | 376 (7.8) | 1,801 (7.5) | 144 (11.8) |
| Osteoarthritis | 2,873 (2.5) | 1,155 (2.6) | 97 (2.2) | 120 (2.6) | 601 (2.6) | 61 (5.5) |
| Fractures (any) | 6,092 (5.2) | 2,298 (5.1) | 212 (4.8) | 231 (4.9) | 1,243 (5.3) | 75 (6.2) |
| Fragility fractures | 2,666 (2.2) | 963 (2.0) | 79 (1.7) | 98 (2.0) | 551 (2.3) | 44 (3.4) |
| Osteoporosis | 1,522 (1.3) | 602 (1.3) | 42 (0.9) | 64 (1.3) | 319 (1.3) | 27 (2.1) |
| Rheumatoid arthritis | 1,131 (0.9) | 479 (1.0) | 38 (0.8) | 33 (0.7) | 238 (1.0) | 15 (1.2) |
| **Brain** | | | | | | |
| Any brain outcome (dementia, delirium, or Parkinson's disease) | 4,619 (3.8) | 1,852 (3.9) | 177 (3.8) | 201 (4.1) | 993 (4.0) | 76 (5.9) |
| Dementia | 2,306 (1.9) | 916 (1.9) | 92 (2.0) | 97 (2.0) | 487 (2.0) | 40 (3.1) |
| Alzheimer's disease | 1,026 (0.8) | 380 (0.8) | 38 (0.8) | 50 (1.0) | 189 (0.8) | 13 (1.0) |
| Non-Alzheimer's dementia | 1,299 (1.1) | 540 (1.1) | 54 (1.2) | 47 (1.0) | 303 (1.2) | 27 (2.1) |
| Delirium | 2,621 (2.1) | 1,028 (2.1) | 95 (2.0) | 112 (2.3) | 603 (2.5) | 45 (3.5) |
| Parkinson's disease | 1,116 (0.9) | 454 (1.0) | 46 (1.0) | 43 (0.9) | 202 (0.8) | 21 (1.6) |
| **Pancreas** | | | | | | |
| T1 or T2 diabetes | 9,515 (8.0) | 3,665 (7.9) | 368 (8.1) | 367 (7.6) | 1,822 (7.7) | 119 (9.6) |
| **Infection** | | | | | | |
| Covid-19 | 2,332 (1.9) | 956 (2.0) | 96 (2.1) | 77 (1.6) | 522 (2.1) | 35 (2.7) |
| Cholecystitis | 1,624 (1.3) | 613 (1.3) | 55 (1.2) | 73 (1.5) | 318 (1.3) | 24 (1.9) |

| | | | | | | |
|---|---|---|---|---|---|---|
| Pneumonia | 8,019 (6.7) | 3,128 (6.7) | 273 (6.0) | 342 (7.1) | 1,651 (6.9) | 95 (7.6) |
| Sepsis | 10,081 (8.4) | 3,955 (8.4) | 374 (8.2) | 416 (8.6) | 2,065 (8.6) | 122 (9.7) |
| LRTI | 4,113 (3.4) | 1,694 (3.6) | 148 (3.2) | 164 (3.3) | 914 (3.7) | 50 (3.9) |
| URTI | 588 (0.5) | 226 (0.5) | 21 (0.5) | 17 (0.3) | 109 (0.4) | 4 (0.3) |
| UTI | 7,099 (5.9) | 2,773 (5.9) | 263 (5.7) | 290 (6.0) | 1,472 (6.1) | 96 (7.6) |
| SSTI | 4,604 (3.8) | 1,792 (3.8) | 176 (3.8) | 216 (4.5) | 1,053 (4.4) | 66 (5.2) |
| **Cardiovascular** | | | | | | |
| Arrhythmia | 2,283 (1.9) | 871 (1.8) | 86 (1.9) | 77 (1.6) | 445 (1.8) | 23 (1.8) |
| Cardiomyopathy | 855 (0.7) | 330 (0.7) | 25 (0.5) | 34 (0.7) | 179 (0.7) | 6 (0.5) |
| CHD | 11,796 (10.5) | 4,626 (10.5) | 412 (9.6) | 485 (10.5) | 2,422 (10.7) | 133 (11.0) |
| Heart failure | 5,779 (4.7) | 2,289 (4.8) | 211 (4.6) | 229 (4.7) | 1,256 (5.1) | 68 (5.3) |
| **Mental Health** | | | | | | |
| Depression | 3,929 (3.4) | 1,615 (3.5) | 132 (3.0) | 152 (3.2) | 772 (3.3) | 46 (3.8) |

Incident disease numbers exclude prevalent disease at baseline. Numbers presented are n (%). Abbreviations: CHD, coronary heart disease; T1, type 1; T2, type 2; LRTI, lower respiratory tract infection; URTI, upper respiratory tract infection; UTI, urinary tract infection; SSTI, skin and soft tissue infection. Joint replacement surgery variable includes a diagnosis of hip, knee, ankle, or shoulder replacement. Any brain outcome variable includes a diagnosis of dementia, delirium, or Parkinson’s disease.

**eTable 5.** Hazard ratios of incident disease outcomes in p.C282Y/H63D genotypes in males

| Males | No mutations | H63D +/- | | H63D +/+ | | C282Y +/ H36D + | | C282Y +/- | | C282Y +/+ | |
|---|---|---|---|---|---|---|---|---|---|---|---|
| | | HR (95% CI) | P | HR (95% CI) | P | HR (95% CI) | P | HR (95% CI) | P | HR (95% CI) | P |
| Hemochromatosis | 1 | 1.53 (1.08-2.17) | 0.02 | 8.95 (5.87-13.64) | $2.20*10^{-24}$ | 27.47 (20.49-36.84) | $1.00*10^{-108}$ | 2.40 (1.66-3.47) | $3.70*10^{-06}$ | 405.29 (317.06-518.11) | $2.97*10^{-501}$ |
| All-cause mortality | 1 | 1.03 (1.00-1.06) | 0.05 | 1.01 (0.93-1.11) | 0.75 | 1.02 (0.94-1.11) | 0.58 | 1.05 (1.00-1.09) | 0.03 | 1.29 (1.12-1.48) | $4.70*10^{-04}$ |
| **Liver** | | | | | | | | | | | |
| Liver disease (any) | 1 | 1.05 (0.99-1.11) | 0.12 | 1.06 (0.90-1.24) | 0.48 | 1.06 (0.91-1.23) | 0.46 | 1.00 (0.93-1.78) | 0.96 | 2.56 (2.10-3.12) | $8.70*10^{-21}$ |
| Alcoholic liver disease | 1 | 0.95 (0.81-1.11) | 0.50 | 1.07 (0.71-1.59) | 0.76 | 1.07 (0.73-1.57) | 0.72 | 1.17 (0.98-1.40) | 0.09 | 1.97 (1.16-3.35) | 0.01 |
| Fibrosis & Cirrhosis | 1 | 1.04 (0.90-1.21) | 0.58 | 1.02 (0.68-1.53) | 0.93 | 1.00 (0.68-1.48) | 0.99 | 1.11 (0.92-1.33) | 0.28 | 5.36 (3.83-7.52) | $2.00*10^{-22}$ |
| Hepatic failure | 1 | 0.99 (0.81-1.21) | 0.92 | 0.74 (0.40-1.40) | 0.36 | 1.04 (0.62-1.74) | 0.88 | 0.75 (0.56-1.00) | 0.05 | 0.81 (0.26-2.52) | 0.72 |
| **Cancer** | | | | | | | | | | | |
| Liver cancer | 1 | 1.17 (0.98-1.41) | 0.09 | 0.86 (0.48-1.53) | 0.61 | 1.32 (0.84-2.08) | 0.22 | 1.07 (0.84-1.36) | 0.61 | 7.90 (5.46-11.43) | $5.50*10^{-28}$ |
| Prostate cancer | 1 | 1.00 (0.96-1.05) | 0.86 | 0.96 (0.86-1.09) | 0.56 | 1.03 (0.93-1.16) | 0.55 | 1.05 (1.00-1.11) | 0.09 | 1.33 (1.09-1.61) | 0.004 |
| **Musculoskeletal** | | | | | | | | | | | |
| Joint replacement surgery (any) | 1 | 1.03 (0.99-1.07) | 0.21 | 0.98 (0.88-1.10) | 0.74 | 1.10 (1.00-1.22) | 0.08 | 1.06 (1.01-1.12) | 0.02 | 1.78 (1.51-2.10) | $6.40*10^{-12}$ |
| Osteoarthritis | 1 | 1.03 (0.96-1.10) | 0.44 | 0.89 (0.72-1.09) | 0.25 | 1.01 (0.84-1.21) | 0.99 | 1.03 (0.94-1.12) | 0.55 | 2.10 (1.63-2.71) | $1.10*10^{-08}$ |
| Fractures (any) | 1 | 0.96 (0.92-1.01) | 0.13 | 0.92 (0.80-1.05) | 0.23 | 0.93 (0.82-1.06) | 0.29 | 1.01 (0.95-1.07) | 0.85 | 1.18 (0.94-1.48) | 0.16 |
| Fragility fractures | 1 | 0.92 (0.86-0.99) | 0.03 | 0.78 (0.62-0.98) | 0.03 | 0.90 (0.74-1.10) | 0.31 | 1.02 (0.93-1.12) | 0.68 | 1.59 (1.18-2.14) | 0.002 |
| Osteoporosis | 1 | 1.01 (0.92-1.11) | 0.85 | 0.73 (0.53-0.99) | 0.04 | 1.03 (0.80-1.32) | 0.82 | 1.02 (0.91-1.16) | 0.70 | 1.70 (1.16-2.48) | 0.007 |
| Rheumatoid arthritis | 1 | 1.08 (0.97-1.21) | 0.13 | 0.89 (0.64-1.23) | 0.48 | 0.73 (0.51-1.03) | 0.07 | 1.05 (0.91-1.20) | 0.53 | 1.30 (0.78-2.16) | 0.31 |
| **Brain** | | | | | | | | | | | |
| Any brain outcome (dementia, delirium, or Parkinson's disease) | 1 | 1.02 (0.97-1.08) | 0.39 | 1.02 (0.88-1.18) | 0.83 | 1.07 (0.93-1.24) | 0.32 | 1.06 (0.99-1.13) | 0.11 | 1.65 (1.31-2.06) | $1.70*10^{-05}$ |
| Dementia | 1 | 1.01 (0.94-1.09) | 0.76 | 1.06 (0.86-1.31) | 0.58 | 1.04 (0.85-1.27) | 0.73 | 1.04 (0.94-1.14) | 0.48 | 1.72 (1.26-2.35) | 0.001 |
| Alzheimer's disease | 1 | 0.94 (0.84-1.06) | 0.32 | 0.98 (0.71-1.36) | 0.91 | 1.21 (0.91-1.60) | 0.19 | 0.90 (0.77-1.05) | 0.20 | 1.26 (0.73-2.17) | 0.42 |
| Non-Alzheimer's dementia | 1 | 1.06 (0.96-1.17) | 0.25 | 1.11 (0.84-1.45) | 0.46 | 0.89 (0.66-1.19) | 0.43 | 1.14 (1.01-1.30) | 0.04 | 2.05 (1.40-3.00) | $2.40*10^{-04}$ |
| Delirium | 1 | 1.00 (0.93-1.08) | 0.98 | 0.96 (0.78-1.18) | 0.69 | 1.06 (0.87-1.28) | 0.57 | 1.13 (1.04-1.24) | 0.006 | 1.69 (1.26-2.27) | $4.80*10^{-04}$ |
| Parkinson's disease | 1 | 1.04 (0.93-1.16) | 0.47 | 1.09 (0.81-1.47) | 0.55 | 0.96 (0.71-1.30) | 0.77 | 0.90 (0.77-1.04) | 0.16 | 1.86 (1.21-2.87) | 0.005 |
| **Pancreas** | | | | | | | | | | | |

| | | | | | | | | | | | |
|---|---|---|---|---|---|---|---|---|---|---|---|
| T1 or T2 diabetes | 1 | 0.98 (0.95-1.02) | 0.44 | 1.02 (0.92-1.13) | 0.69 | 0.95 (0.86-1.06) | 0.35 | 0.95 (0.91-1.00) | 0.06 | 1.25 (1.05-1.50) | 0.01 |
| **Infection** | | | | | | | | | | | |
| COVID-19 | 1 | 1.05 (0.97-1.13) | 0.21 | 1.09 (0.89-1.34) | 0.4 | 0.81 (0.65-1.02) | 0.07 | 1.11 (1.01-1.22) | 0.03 | 1.51 (1.08-2.11) | 0.02 |
| Cholecystitis | 1 | 0.96 (0.88-1.06) | 0.45 | 0.89 (0.68-1.16) | 0.39 | 1.11 (0.87-1.40) | 0.4 | 0.96 (0.86-1.09) | 0.56 | 1.41 (0.94-2.11) | 0.09 |
| Pneumonia | 1 | 1.00 (0.96-1.04) | 0.82 | 0.90 (0.80-1.02) | 0.09 | 1.04 (0.93-1.16) | 0.51 | 1.01 (0.96-1.07) | 0.68 | 1.14 (0.93-1.40) | 0.21 |
| Sepsis | 1 | 1.01 (0.97-1.04) | 0.78 | 0.98 (0.88-1.08) | 0.66 | 1.02 (0.92-1.12) | 0.7 | 1.01 (0.96-1.06) | 0.70 | 1.17 (0.98-1.40) | 0.09 |
| LRTI | 1 | 1.05 (0.99-1.11) | 0.10 | 0.94 (0.80-1.11) | 0.46 | 0.96 (0.82-1.12) | 0.58 | 1.08 (1.00-1.16) | 0.48 | 1.11 (0.84-1.46) | 0.48 |
| URTI | 1 | 0.99 (0.84-1.15) | 0.85 | 0.94 (0.61-1.45) | 0.77 | 0.71 (0.44-1.15) | 0.16 | 0.92 (0.75-1.13) | 0.41 | 0.64 (0.24-1.71) | 0.38 |
| UTI | 1 | 1.00 (0.95-1.04) | 0.90 | 0.98 (0.86-1.10) | 0.69 | 1.00 (0.89-1.13) | 0.98 | 1.02 (0.96-1.08) | 0.51 | 1.31 (1.07-1.61) | 0.008 |
| SSTI | 1 | 1.00 (0.94-1.05) | 0.88 | 1.01 (0.87-1.17) | 0.90 | 1.16 (1.01-1.33) | 0.03 | 1.14 (1.06-1.22) | $1.80*10^{-04}$ | 1.39 (1.09-1.77) | 0.008 |
| **Cardiovascular** | | | | | | | | | | | |
| Arrhythmia | 1 | 0.98 (0.90-1.06) | 0.57 | 1.00 (0.81-1.24) | 0.99 | 0.84 (0.67-1.05) | 0.13 | 0.96 (0.87-1.07) | 0.49 | 0.97 (0.64-1.47) | 0.89 |
| Cardiomyopathy | 1 | 0.99 (0.87-1.12) | 0.88 | 0.77 (0.52-1.15) | 0.21 | 0.99 (0.70-1.39) | 0.94 | 1.04 (0.89-1.23) | 0.60 | 0.69 (0.31-1.53) | 0.36 |
| CHD | 1 | 1.01 (0.97-1.04) | 0.77 | 0.91 (0.83-1.01) | 0.08 | 1.00 (0.91-1.10) | 0.99 | 1.02 (0.97-1.06) | 0.45 | 1.06 (0.90-1.26) | 0.49 |
| Heart failure | 1 | 1.01 (0.96-1.06) | 0.67 | 0.96 (0.84-1.10) | 0.59 | 0.97 (0.85-1.11) | 0.65 | 1.07 (1.01-1.14) | 0.03 | 1.14 (0.89-1.44) | 0.29 |
| **Mental Health** | | | | | | | | | | | |
| Depression | 1 | 1.05 (0.99-1.12) | 0.07 | 0.88 (0.74-1.05) | 0.16 | 0.96 (0.81-1.13) | 0.60 | 0.98 (0.91-1.06) | 0.59 | 1.15 (0.86-1.54) | 0.34 |

HR (Hazard ratio) compared to those with neither *HFE* mutation. Cox proportional hazards regression models adjusted for age, assessment centre, and genetic principal components 1–10. Abbreviations: CHD, coronary heart disease; T1, type 1; T2, type 2; LRTI, lower respiratory tract infection; URTI, upper respiratory tract infection; UTI, urinary tract infection; SSTI, skin and soft tissue infection; CI, confidence interval. Joint replacement surgery variable includes a diagnosis of hip, knee, ankle, or shoulder replacement. Any brain outcome variable includes a diagnosis of dementia, delirium, or Parkinson's disease.

**eTable 6**. Incident hospital diagnoses in male UK Biobank participants by p.C282Y/H63D genotypes, excluding a diagnosis of hemochromatosis at baseline

| **Males** | **No mutations** | **H63D +/-** | **H63D +/+** | **C282Y +/ H36D +** | **C282Y +/-** | **C282Y +/+** |
|---|---|---|---|---|---|---|
| Hemochromatosis | 85 (14.41) | 51 (8.64) | 29 (4.92) | 95 (16.10) | 42 (7.12) | 288 (48.81) |
| All-cause mortality | 13959 (58.44) | 5653 (23.66) | 539 (2.26) | 587 (2.46) | 2990 (12.52) | 160 (0.67) |
| **Liver** | | | | | | |
| Liver disease (any) | 3969 (58.38) | 1619 (23.82) | 158 (2.32) | 167 (2.46) | 800 (11.77) | 85 (1.25) |
| Alcoholic liver disease | 607 (58.20) | 226 (21.67) | 25 (2.40) | 27 (2.59) | 148 (14.19) | 10 (0.96) |
| Fibrosis & Cirrhosis | 618 (57.06) | 250 (23.08) | 23 (2.12) | 25 (2.31) | 140 (12.93) | 27 (2.49) |
| Hepatic failure | 355 (62.06) | 137 (23.95) | 10 (1.75) | 15 (2.65) | 54 (9.44) | 1 (0.17) |
| **Cancer** | | | | | | |
| Liver cancer | 361 (55.28) | 166 (25.42) | 12 (1.84) | 19 (2.91) | 78 (11.94) | 17 (2.60) |
| Prostate cancer | 7722 (59.20) | 3023 (23.18) | 283 (2.17) | 318 (2.44) | 1610 (12.34) | 87 (0.67) |
| **Musculoskeletal** | | | | | | |
| Joint replacement surgery (any) | 8427 (58.44) | 3377 (23.42) | 315 (2.18) | 375 (2.60) | 1801 (12.49) | 126 (0.87) |
| Osteoarthritis | 2872 (58.70) | 1153 (23.56) | 97 (1.98) | 120 (2.45) | 600 (12.26) | 51 (1.04) |
| Fractures (any) | 6090 (60.07) | 2296 (22.65) | 212 (2.09) | 231 (2.28) | 1243 (12.26) | 66 (0.65) |
| Fragility fractures | 2663 (60.63) | 963 (21.93) | 79 (1.80) | 98 (2.23) | 550 (12.52) | 39 (0.89) |
| Osteoporosis | 1521 (59.37) | 601 (23.46) | 42 (1.64) | 63 (2.46) | 319 (12.45) | 16 (0.62) |
| Rheumatoid arthritis | 1130 (58.55) | 479 (24.82) | 38 (1.97) | 33 (1.71) | 238 (12.33) | 12 (0.62) |
| **Brain** | | | | | | |
| Any brain outcome (dementia, delirium, or Parkinson's disease) | 4616 (58.43) | 1849 (23.41) | 177 (2.24) | 200 (2.53) | 992 (12.56) | 66 (0.84) |
| Dementia | 2305 (58.67) | 915 (23.29) | 92 (2.34) | 97 (2.47) | 487 (12.40) | 33 (0.84) |
| Alzheimer's disease | 1026 (60.53) | 380 (22.42) | 38 (2.24) | 50 (2.95) | 189 (11.15) | 12 (0.71) |
| Non-Alzheimer's dementia | 1298 (57.38) | 539 (23.83) | 54 (2.39) | 47 (2.08) | 303 (13.40) | 21 (0.93) |
| Delirium | 2619 (58.29) | 1025 (22.81) | 95 (2.11) | 111 (2.47) | 602 (13.40) | 41 (0.91) |
| Parkinson's disease | 1115 (59.34) | 454 (24.16) | 46 (2.45) | 43 (2.29) | 202 (10.75) | 19 (1.01) |
| **Pancreas** | | | | | | |

| | | | | | | |
|---|---|---|---|---|---|---|
| T2 diabetes | 9570 (60.17) | 3673 (23.09) | 365 (2.30) | 368 (2.31) | 1831 (11.51) | 97 (0.61) |
| **Infection** | | | | | | |
| Covid-19 | 2331 (58.14) | 953 (23.77) | 96 (2.39) | 76 (1.90) | 522 (13.02) | 31 (0.77) |
| Cholecystitis | 1623 (60.07) | 612 (22.65) | 55 (2.04) | 73 (2.70) | 316 (11.70) | 23 (0.85) |
| Pneumonia | 8014 (59.44) | 3124 (23.17) | 273 (2.02) | 340 (2.52) | 1649 (12.23) | 83 (0.62) |
| Sepsis | 10075 (59.34) | 3951 (23.27) | 374 (2.20) | 412 (2.43) | 2063(12.15) | 103 (0.61) |
| LRTI | 4110 (58.12) | 1691 (23.91) | 148 (2.09) | 164 (2.32) | 913 (12.91) | 45 (0.64) |
| URTI | 588 (61.06) | 226 (23.47) | 21 (2.18) | 17 (1.77) | 108 (11.21) | 3 (0.31) |
| UTI | 7097 (59.29) | 2769 (23.13) | 263 (2.20) | 286 (2.39) | 1470 (12.28) | 85 (0.71) |
| SSTI | 4599 (58.27) | 1790 (22.68) | 176 (2.23) | 215 (2.72) | 1053 (13.34) | 59 (0.75) |
| **Cardiovascular** | | | | | | |
| Arrhythmia | 2282 (60.40) | 870 (23.03) | 85 (2.25) | 77 (2.04) | 444 (11.75) | 20 (0.53) |
| Cardiomyopathy | 853 (59.86) | 330 (23.16) | 25 (1.75) | 34 (2.39) | 178 (12.49) | 5 (0.35) |
| CHD | 11793 (59.45) | 4622 (23.30) | 411 (2.07) | 481 (2.42) | 2414 (12.17) | 115 (0.58) |
| Heart failure | 5775 (58.87) | 2285 (23.29) | 211 (2.15) | 227 (2.31) | 1253 (12.77) | 59 (0.60) |
| **Mental Health** | | | | | | |
| Depression | 3925 (59.18) | 1614 (24.34) | 132 (1.99) | 150 (2.26) | 771 (11.63) | 40 (0.60) |

Incident disease numbers exclude prevalent disease at baseline. Numbers presented are n (%). Abbreviations: CHD, coronary heart disease; T1, type 1; T2, type 2; LRTI, lower respiratory tract infection; URTI, upper respiratory tract infection; UTI, urinary tract infection; SSTI, skin and soft tissue infection. Joint replacement surgery variable includes a diagnosis of hip, knee, ankle, or shoulder replacement. Any brain outcome variable includes a diagnosis of dementia, delirium, or Parkinson's disease.

**eTable 7.** Hazard ratios of incident disease outcomes in p.C282Y/H63D genotypes in males, excluding a diagnosis of hemochromatosis at baseline

| Males | No mutations (Ref group) | H63D +/- | | H63D +/+ | | C282Y +/ H36D + | | C282Y +/- | | C282Y +/+ | |
|---|---|---|---|---|---|---|---|---|---|---|---|
| | | HR (95% CI) | P | HR (95% CI) | P | HR (95% CI) | P | HR (95% CI) | P | HR (95% CI) | P |
| Hemochromatosis | 1 | 1.53 (1.08-2.17) | 0.02 | 8.95 (5.87-13.64) | $2.2*10^{-24}$ | 27.49 (20.50-36.86) | $1.0*10^{-108}$ | 2.40 (1.66-3.47) | $3.6*10^{-06}$ | 405.79 (317.34-518.75) | 0.00E+00 |
| All-cause mortality | 1 | 1.03 (1.00-1.07) | 0.04 | 1.01 (0.93-1.11) | 0.74 | 1.02 (0.94-1.11) | 0.58 | 1.05 (1.00-1.09) | 0.03 | 1.22 (1.05-1.43) | 0.01 |
| **Liver** | | | | | | | | | | | |
| Liver disease (any) | 1 | 1.05 (0.99-1.11) | 0.13 | 1.05 (0.89-1.23) | 0.57 | 1.04 (0.89-1.21) | 0.63 | 1.00 (0.92-1.07) | 0.9 | 2.36 (1.90-2.93) | $5.3*10^{-15}$ |
| Alcoholic liver disease | 1 | 0.95 (0.12-1.10) | 0.48 | 1.07 (0.72-1.60) | 0.74 | 1.04 (0.71-1.53) | 0.84 | 1.15 (0.96-1.38) | 0.12 | 1.59 (0.85-2.97) | 0.15 |
| Fibrosis & Cirrhosis | 1 | 1.03 (0.89-1.20) | 0.66 | 0.98 (0.64-1.48) | 0.91 | 0.97 (0.65-1.44) | 0.87 | 1.09 (0.91-1.31) | 0.35 | 4.52 (3.07-6.66) | $2.1*10^{-14}$ |
| Hepatic failure | 1 | 0.99 (0.81-1.21) | 0.92 | 0.75 (0.40-1.40) | 0.36 | 1.05 (0.62-1.76) | 0.86 | 0.75 (0.56-1.00) | 0.05 | 0.31 (0.04-2.19) | 0.24 |
| **Cancer** | | | | | | | | | | | |
| Liver cancer | 1 | 1.17 (0.98-1.41) | 0.09 | 0.87 (0.49-1.54) | 0.63 | 1.27 (0.80-2.02) | 0.3 | 1.05 (0.82-1.34) | 0.71 | 4.97 (3.05-8.11) | $1.2*10^{-10}$ |
| Prostate cancer | 1 | 1.00 (0.96-1.05) | 0.87 | 0.97 (0.86-1.09) | 0.57 | 1.04 (0.93-1.16) | 0.51 | 1.05 (0.99-1.11) | 0.09 | 1.27 (1.03-1.57) | 0.03 |
| **Musculoskeletal** | | | | | | | | | | | |
| Joint replacement surgery (any) | 1 | 1.03 (0.99-1.07) | 0.22 | 0.98 (0.88-1.10) | 0.75 | 1.10 (0.99-1.22) | 0.06 | 1.06 (1.01-1.12) | 0.02 | 1.75 (1.47-2.09) | $4.8*10^{-10}$ |
| Osteoarthritis | 1 | 1.03 (0.96-1.10) | 0.46 | 0.89 (0.73-1.09) | 0.26 | 1.02 (0.85-1.22) | 0.87 | 1.03 (0.94-1.12) | 0.56 | 1.97 (1.50-2.60) | $1.5*10^{-06}$ |
| Fractures (any) | 1 | 0.96 (0.92-1.01) | 0.12 | 0.92 (0.80-1.06) | 0.24 | 0.94 (0.82-1.07) | 0.34 | 1.01 (0.95-1.07) | 0.82 | 1.17 (0.91-1.49) | 0.22 |
| Fragility fractures | 1 | 0.92 (0.86-0.99) | 0.03 | 0.78 (0.63-0.98) | 0.03 | 0.91 (0.74-1.11) | 0.34 | 1.02 (0.93-1.12) | 0.68 | 1.60 (1.17-2.20) | $3.6*10^{-03}$ |
| Osteoporosis | 1 | 1.01 (0.92-1.11) | 0.86 | 0.73 (0.54-0.99) | 0.04 | 1.02 (0.79-1.31) | 0.89 | 1.03 (0.91-1.16) | 0.68 | 1.14 (0.69-1.86) | 0.61 |
| Rheumatoid arthritis | 1 | 1.09 (0.98-1.21) | 0.13 | 0.89 (0.64-1.23) | 0.48 | 0.73 (0.52-1.03) | 0.07 | 1.05 (0.91-1.20) | 0.52 | 1.17 (0.66-2.07) | 0.58 |
| **Brain** | | | | | | | | | | | |
| Any brain outcome (dementia, delirium, or Parkinson's disease) | 1 | 1.02 (0.97-1.08) | 0.4 | 1.02 (0.88-1.18) | 0.8 | 1.08 (0.93-1.24) | 0.31 | 1.06 (0.99-1.13) | 0.1 | 1.62 (1.27-2.07) | $1.0*10^{-04}$ |
| Dementia | 1 | 1.01 (0.94-1.09) | 0.76 | 1.06 (0.86-1.31) | 0.57 | 1.04 (0.85-1.28) | 0.69 | 1.04 (0.94-1.14) | 0.46 | 1.60 (1.14-2.26) | 0.01 |
| Alzheimer's disease | 1 | 0.94 (0.84-1.06) | 0.32 | 0.98 (0.71-1.36) | 0.92 | 1.12 (0.91-1.61) | 0.18 | 0.90 (0.77-1.06) | 0.2 | 1.31 (0.74-2.31) | 0.35 |
| Non-Alzheimer's dementia | 1 | 1.06 (0.96-1.17) | 0.26 | 1.11 (0.85-1.46) | 0.45 | 0.89 (0.67-1.20) | 0.45 | 1.14 (1.01-1.30) | 0.04 | 1.81 (1.17-2.78) | 0.01 |
| Delirium | 1 | 1.00 (0.93-1.07) | 0.98 | 0.96 (0.78-1.18) | 0.7 | 1.05 (0.87-1.27) | 0.59 | 1.13 (1.04-1.24) | 0.01 | 1.75 (1.28-2.38) | $3.9*10^{-04}$ |
| Parkinson's disease | 1 | 1.07 (0.98-1.16) | 0.13 | 1.05 (0.84-1.31) | 0.67 | 1.06 (0.85-1.33) | 0.58 | 1.01 (0.91-1.13) | 0.79 | 1.44 (1.02-2.03) | 0.04 |

| | | | | | | | | | | | |
|---|---|---|---|---|---|---|---|---|---|---|---|
| **Pancreas** | | | | | | | | | | | |
| T1 or T2 diabetes | 1 | 0.99 (0.95-1.02) | 0.45 | 1.02 (0.92-1.14) | 0.66 | 0.94 (0.85-1.05) | 0.28 | 0.95 (0.91-1.00) | 0.06 | 1.17 (0.96-1.43) | 0.11 |
| **Infection** | | | | | | | | | | | |
| COVID-19 | 1 | 1.05 (0.97-1.13) | 0.23 | 1.09 (0.89-1.34) | 0.39 | 0.81 (0.64-1.01) | 0.07 | 1.11 (1.01-1.23) | 0.03 | 1.51 (1.06-2.15) | 0.02 |
| Cholecystitis | 1 | 0.96 (0.88-1.06) | 0.44 | 0.89 (0.68-1.17) | 0.4 | 1.11 (0.88-1.41) | 0.37 | 0.96 (0.85-1.08) | 0.51 | 1.54 (1.02-2.32) | 0.04 |
| Pneumonia | 1 | 0.99 (0.95-1.04) | 0.8 | 0.90 (0.80-1.02) | 0.09 | 1.04 (0.93-1.16) | 0.52 | 1.01 (0.96-1.07) | 0.66 | 1.13 (0.91-1.40) | 0.27 |
| Sepsis | 1 | 1.01 (0.97-1.04) | 0.79 | 0.98 (0.88-1.09) | 0.69 | 1.01 (0.92-1.12) | 0.82 | 1.01 (0.96-1.06) | 0.68 | 1.12 (0.92-1.36) | 0.26 |
| LRTI | 1 | 1.05 (0.99-1.11) | 0.1 | 0.94 (0.80-1.11) | 0.48 | 0.96 (0.82-1.13) | 0.63 | 1.08 (1.00-1.16) | 0.04 | 1.14 (0.85-1.52) | 0.4 |
| URTI | 1 | 0.99 (0.85-1.15) | 0.85 | 0.94 (0.61-1.45) | 0.78 | 0.71 (0.44-1.16) | 0.17 | 0.91 (0.74-1.12) | 0.37 | 0.55 (0.18-1.70) | 0.3 |
| UTI | 1 | 1.00 (0.95-1.04) | 0.86 | 0.98 (0.86-1.11) | 0.71 | 1.00 (0.89-1.12) | 0.96 | 1.02 (0.96-1.08) | 0.51 | 1.32 (1.07-1.64) | 0.01 |
| SSTI | 1 | 1.00 (0.94-1.05) | 0.89 | 1.01 (0.87-1.18) | 0.87 | 1.16 (1.01-1.33) | 0.03 | 1.14 (1.07-1.22) | $1.4*10^{-04}$ | 1.40 (1.09-1.82) | 0.01 |
| **Cardiovascular** | | | | | | | | | | | |
| Arrhythmia | 1 | 0.98 (0.90-1.06) | 0.56 | 0.99 (0.80-1.23) | 0.92 | 0.84 (0.67-1.06) | 0.15 | 0.96 (0.87-1.07) | 0.48 | 0.96 (0.62-1.49) | 0.86 |
| Cardiomyopathy | 1 | 0.99 (0.87-1.13) | 0.91 | 0.78 (0.52-1.16) | 0.21 | 0.99 (0.71-1.40) | 0.98 | 1.04 (0.89-1.22) | 0.62 | 0.65 (0.27-1.57) | 0.34 |
| CHD | 1 | 1.00 (0.97-1.04) | 0.8 | 0.91 (0.83-1.01) | 0.71 | 1.00 (0.91-1.09) | 0.97 | 1.01 (0.97-1.06) | 0.52 | 1.04 (0.87-1.25) | 0.66 |
| Heart failure | 1 | 1.01 (0.96-1.06) | 0.69 | 0.96 (0.84-1.11) | 0.61 | 0.97 (0.85-1.10) | 0.62 | 1.07 (1.01-1.14) | 0.03 | 1.12 (0.87-1.45) | 0.38 |
| **Mental Health** | | | | | | | | | | | |
| Depression | 1 | 1.05 (1.00-1.12) | 0.07 | 0.88 (0.74-1.05) | 0.17 | 0.95 (0.81-1.12) | 0.55 | 0.98 (0.91-1.06) | 0.6 | 1.13 (0.82-1.54) | 0.46 |

HR (Hazard ratio) compared to those with neither *HFE* mutation. Cox proportional hazards regression models adjusted for age, assessment centre, and genetic principal components 1–10. Abbreviations: CHD, coronary heart disease; T1, type 1; T2, type 2; LRTI, lower respiratory tract infection; URTI, upper respiratory tract infection; UTI, urinary tract infection SSTI, skin and soft tissue infection; CI, confidence interval. Joint replacement surgery variable includes a diagnosis of hip, knee, ankle, or shoulder replacement. Any brain outcome variable includes a diagnosis of dementia, delirium, or Parkinson's disease.

**eTable 8**. Cumulative incidence of hemochromatosis from ages 40-80 years by HFE genotypes

| | **Total Cohort (n=451,270)** | | | **No mutations** | | | **H63D Heterozygotes** | | | **H63D Homozygotes** | | | **Compound C282Y/H63D Heterozygotes** | | | **p.C282Y Heterozygotes** | | | **p.C282Y homozygotes** | | |
|---|---|---|---|---|---|---|---|---|---|---|---|---|---|---|---|---|---|---|---|---|---|
| | **Incident diagnosis** | | | **Incident diagnosis** | | | **Incident diagnosis** | | | **Incident diagnosis** | | | **Incident diagnosis** | | | **Incident diagnosis** | | | **Incident diagnosis** | | |
| | **%** | **(95% CI)** | | **%** | **(95% CI)** | | **%** | **(95% CI)** | | **%** | **(95% CI)** | | **%** | **(95% CI)** | | **%** | **(95% CI)** | | **%** | **(95% CI)** | |
| **Male age group (years)** | | | | | | | | | | | | | | | | | | | | | |
| **40 - 45** | 0.1 | 0.0 | 0.2 | 0.0 | 0.0 | 0.2 | 0.0 | 0.0 | 0.0 | 0.0 | 0.0 | 0.0 | 0.3 | 0.0 | 1.9 | 0.0 | 0.0 | 0.0 | 6.8 | 2.8 | 16.1 |
| **46 - 50** | 0.1 | 0.1 | 0.2 | 0.0 | 0.0 | 0.2 | 0.0 | 0.0 | 0.1 | 0.4 | 0.1 | 1.3 | 0.7 | 0.2 | 2.0 | 0.0 | 0.0 | 0.0 | 12.5 | 7.3 | 20.8 |
| **51 - 55** | 0.2 | 0.2 | 0.3 | 0.0 | 0.0 | 0.2 | 0.0 | 0.0 | 0.1 | 0.6 | 0.3 | 1.5 | 1.5 | 0.8 | 2.6 | 0.0 | 0.0 | 0.1 | 20.4 | 14.7 | 27.9 |
| **56 - 60** | 0.3 | 0.3 | 0.4 | 0.1 | 0.0 | 0.2 | 0.1 | 0.0 | 0.1 | 0.8 | 0.4 | 1.6 | 2.1 | 1.3 | 3.2 | 0.1 | 0.1 | 0.2 | 28.4 | 22.7 | 35.3 |
| **61 - 65** | 0.4 | 0.3 | 0.5 | 0.1 | 0.0 | 0.2 | 0.1 | 0.1 | 0.2 | 1.0 | 0.6 | 1.8 | 2.6 | 1.8 | 3.8 | 0.2 | 0.1 | 0.3 | 36.1 | 30.5 | 42.4 |
| **66 - 70** | 0.5 | 0.5 | 0.6 | 0.1 | 0.1 | 0.2 | 0.2 | 0.1 | 0.2 | 1.2 | 0.7 | 2.1 | 3.6 | 2.7 | 4.8 | 0.3 | 0.2 | 0.4 | 43.0 | 37.6 | 48.7 |
| **71 - 75** | 0.6 | 0.6 | 0.7 | 0.2 | 0.1 | 0.2 | 0.2 | 0.1 | 0.3 | 1.4 | 0.9 | 2.2 | 4.3 | 3.4 | 5.5 | 0.3 | 0.2 | 0.4 | 48.9 | 43.8 | 54.2 |
| **76 - 80** | 0.8 | 0.7 | 0.9 | 0.2 | 0.1 | 0.3 | 0.3 | 0.2 | 0.4 | 1.9 | 1.3 | 2.9 | 5.4 | 4.3 | 6.8 | 0.4 | 0.3 | 0.6 | 56.4 | 51.4 | 61.6 |
| **Female age group (years)** | | | | | | | | | | | | | | | | | | | | | |
| **40 - 45** | 0.0 | 0.0 | 0.0 | 0.0 | 0.0 | 0.0 | 0.0 | 0.0 | 0.0 | 0.0 | 0.0 | 0.0 | 0.2 | 0.0 | 1.5 | 0.0 | 0.0 | 0.0 | 0.0 | 0.0 | 0.0 |
| **46 - 50** | 0.0 | 0.0 | 0.1 | 0.0 | 0.0 | 0.0 | 0.0 | 0.0 | 0.1 | 0.2 | 0.1 | 0.9 | 0.3 | 0.1 | 1.3 | 0.0 | 0.0 | 0.0 | 3.4 | 1.7 | 6.7 |
| **51 - 55** | 0.1 | 0.1 | 0.1 | 0.0 | 0.0 | 0.0 | 0.0 | 0.0 | 0.1 | 0.3 | 0.1 | 1.0 | 0.5 | 0.2 | 1.3 | 0.0 | 0.0 | 0.1 | 8.2 | 5.7 | 11.7 |
| **56 - 60** | 0.1 | 0.1 | 0.2 | 0.0 | 0.0 | 0.0 | 0.1 | 0.0 | 0.1 | 0.4 | 0.2 | 1.0 | 0.7 | 0.4 | 1.5 | 0.1 | 0.1 | 0.2 | 14.5 | 11.5 | 18.1 |
| **61 - 65** | 0.2 | 0.2 | 0.3 | 0.0 | 0.0 | 0.0 | 0.1 | 0.0 | 0.1 | 0.5 | 0.2 | 1.1 | 1.1 | 0.6 | 1.8 | 0.1 | 0.1 | 0.2 | 21.2 | 17.9 | 24.9 |
| **66 - 70** | 0.3 | 0.3 | 0.3 | 0.0 | 0.0 | 0.1 | 0.1 | 0.1 | 0.2 | 0.5 | 0.2 | 1.1 | 1.5 | 1.0 | 2.2 | 0.2 | 0.1 | 0.3 | 27.6 | 24.2 | 31.3 |
| **71 - 75** | 0.4 | 0.3 | 0.4 | 0.1 | 0.0 | 0.1 | 0.1 | 0.1 | 0.2 | 0.6 | 0.3 | 1.2 | 1.9 | 1.4 | 2.7 | 0.2 | 0.2 | 0.3 | 33.5 | 30.0 | 37.2 |
| **76 - 80** | 0.5 | 0.5 | 0.6 | 0.1 | 0.1 | 0.1 | 0.1 | 0.1 | 0.2 | 0.6 | 0.3 | 1.3 | 2.7 | 2.0 | 3.6 | 0.3 | 0.2 | 0.5 | 40.5 | 36.7 | 44.5 |

**eTable 9**. Incident hospital diagnoses in female UK Biobank participants by p.C282Y/H63D genotypes

| Females | No mutations | H63D +/- | H63D +/+ | C282Y +/ H36D + | C282Y +/- | C282Y +/+ |
|---|---|---|---|---|---|---|
| Hemochromatosis | 44 (0.03) | 35 (0.1) | 12 (0.2) | 57 (1.0) | 37 (0.1) | 291 (18.8) |
| All-cause mortality | 9,849 (6.8) | 3,810 (6.7) | 363 (6.5) | 393 (6.8) | 2,020 (6.9) | 129 (8.0) |
| **Liver** | | | | | | |
| Liver disease (any) | 3,822 (2.6) | 1,512 (2.7) | 146 (2.6) | 179 (3.1) | 825 (2.9) | 69 (4.3) |
| Alcoholic liver disease | 181 (0.1) | 78 (0.1) | 11 (0.2) | 6 (0.1) | 59 (0.2) | 7 (0.4) |
| Fibrosis & Cirrhosis | 475 (0.3) | 163 (0.3) | 16 (0.3) | 21 (0.4) | 89 (0.3) | 14 (0.9) |
| Hepatic failure | 200 (0.1) | 60 (0.1) | 7 (0.1) | 4 (0.1) | 35 (0.1) | 5 (0.3) |
| **Cancer** | | | | | | |
| Liver cancer | 232 (0.2) | 114 (0.2) | 10 (0.2) | 8 (0.1) | 49 (0.2) | 3 (0.2) |
| **Musculoskeletal** | | | | | | |
| Joint replacement surgery (any) | 12,356 (8.7) | 4,772 (8.6) | 454 (8.4) | 501 (8.9) | 2,415 (8.5) | 158 (10.3) |
| Osteoarthritis | 4,344 (3.3) | 1,681 (3.3) | 177 (3.6) | 190 (3.7) | 893 (3.4) | 70 (5.1) |
| Fractures (any) | 10,666 (7.6) | 4,204 (7.7) | 376 (7.0) | 430 (7.7) | 2,189 (7.8) | 124 (8.0) |
| Fragility fractures | 6,308 (4.4) | 2,517 (4.5) | 214 (3.9) | 275 (4.8) | 1,265 (4.4) | 63 (4.0) |
| Osteoporosis | 6,990 (5.0) | 2,580 (4.7) | 254 (4.7) | 245 (4.4) | 1,390 (4.9) | 103 (6.7) |
| Rheumatoid arthritis | 2,189 (1.5) | 819 (1.5) | 79 (1.4) | 87 (1.5) | 391 (1.4) | 26 (1.6) |
| **Brain** | | | | | | |
| Any brain outcome (dementia, delirium, or Parkinson's disease) | 3,876 (2.7) | 1,547 (2.7) | 142 (2.6) | 155 (2.7) | 777 (2.7) | 61 (3.8) |
| Dementia | 2,159 (1.5) | 837 (1.5) | 68 (1.2) | 84 (1.5) | 440 (1.5) | 31 (1.9) |
| Alzheimer's disease | 1,072 (0.7) | 450 (0.8) | 37 (0.7) | 51 (0.9) | 220 (0.8) | 14 (0.9) |
| Non-Alzheimer's dementia | 1,099 (0.8) | 395 (0.7) | 31 (0.6) | 34 (0.6) | 227 (0.8) | 17 (1.1) |
| Delirium | 1,983 (1.4) | 823 (1.4) | 81 (1.5) | 82 (1.4) | 406 (1.4) | 36 (2.2) |
| Parkinson's disease | 691 (0.5) | 249 (0.4) | 26 (0.5) | 30 (0.5) | 134 (0.5) | 8 (0.5) |
| **Pancreas** | | | | | | |
| T1 or T2 diabetes | 6,532 (4.6) | 2,583 (4.6) | 245 (4.5) | 278 (4.9) | 1,327 (4.6) | 87 (5.5) |
| **Infection** | | | | | | |
| Covid-19 | 1,809 (1.2) | 720 (1.3) | 72 (1.3) | 72 (1.3) | 383 (1.3) | 21 (1.3) |
| Cholecystitis | 1,749 (1.2) | 675 (1.2) | 62 (1.1) | 69 (1.2) | 351 (1.2) | 30 (1.9) |
| Pneumonia | 6,130 (4.3) | 2,408 (4.3) | 242 (4.4) | 255 (4.5) | 1,276 (4.5) | 75 (4.8) |

| | | | | | | |
|---|---|---|---|---|---|---|
| Sepsis | 10,026 (7.0) | 3,840 (6.9) | 379 (7.0) | 414 (7.4) | 2,103 (7.4) | 133 (8.5) |
| LRTI | 3,875 (2.7) | 1,439 (2.5) | 139 (2.5) | 144 (2.5) | 821 (2.8) | 53 (3.3) |
| URTI | 636 (0.4) | 279 (0.5) | 21 (0.4) | 21 (0.4) | 142 (0.5) | 2 (0.1) |
| UTI | 9,140 (6.5) | 3,493 (6.4) | 351 (6.6) | 366 (6.7) | 1,905 (6.8) | 126 (8.2) |
| SSTI | 3,900 (2.7) | 1,534 (2.7) | 158 (2.9) | 153 (2.7) | 799 (2.8) | 53 (3.3) |
| **Cardiovascular** | | | | | | |
| Arrhythmia | 1,655 (1.1) | 656 (1.2) | 48 (0.9) | 67 (1.2) | 323 (1.1) | 20 (1.3) |
| Cardiomyopathy | 608 (0.4) | 230 (0.4) | 19 (0.3) | 26 (0.5) | 130 (0.5) | 3 (0.2) |
| CHD | 7,402 (5.3) | 2,877 (5.2) | 294 (5.4) | 293 (5.3) | 1,502 (5.3) | 82 (5.3) |
| Heart failure | 3,551 (2.4) | 1,431 (2.5) | 127 (2.3) | 155 (2.7) | 753 (2.6) | 56 (3.5) |
| **Mental Health** | | | | | | |
| Depression | 6,415 (4.8) | 2430 (4.6) | 269 (5.2) | 279 (5.2) | 1317 (4.9) | 64 (4.3) |

Incident disease numbers exclude prevalent disease at baseline. Numbers presented are n (%). Abbreviations: CHD, coronary heart disease; T1, type 1; T2, type 2; LRTI, lower respiratory tract infection; URTI, upper respiratory tract infection; UTI, urinary tract infection; SSTI, skin and soft tissue infection. Joint replacement surgery variable includes a diagnosis of hip, knee, ankle, or shoulder replacement. Any brain outcome variable includes a diagnosis of dementia, delirium, or Parkinson’s disease.

**eTable 10.** Hazard ratios of incident disease outcomes in p.C282Y/H63D genotypes in females

| Females | No mutations | H63D +/- | | H63D +/+ | | C282Y +/ H36D + | | C282Y +/- | | C282Y +/+ | |
|---|---|---|---|---|---|---|---|---|---|---|---|
| | | HR (95% CI) | P | HR (95% CI) | P | HR (95% CI) | P | HR (95% CI) | P | HR (95% CI) | P |
| Hemochromatosis | 1 | 2.04 (1.31-3.17) | 1.70*10-03 | 7.10 (3.75-13.45) | 1.80*10-09 | 32.78 (22.10-48.63) | 2.20*10-67 | 4.15 (2.68-6.44) | 1.80*10-10 | 674.10 (489.11-929.05) | 2.47*10-346 |
| All-cause mortality | 1 | 0.99 (0.96-1.03) | 0.68 | 0.95 (0.85-1.05) | 0.3 | 1.01 (0.91-1.11) | 0.9 | 1.01 (0.96-1.06) | 0.62 | 1.10 (0.92-1.31) | 0.28 |
| **Liver** | | | | | | | | | | | |
| Liver disease (any) | 1 | 1.01 (0.95-1.07) | 0.72 | 1.00 (0.84-1.17) | 0.96 | 1.19 (1.02-1.38) | 0.03 | 1.08 (1.00-1.16) | 0.06 | 1.62 (1.27-2.05) | 7.80*10-05 |
| Alcoholic liver disease | 1 | 1.09 (0.84-1.43) | 0.51 | 1.56 (0.85-2.87) | 0.15 | 0.78 (0.35-1.77) | 0.56 | 1.53 (1.14-2.05) | 0.005 | 3.07 (1.44-6.54) | 0.004 |
| Fibrosis & Cirrhosis | 1 | 0.87 (0.73-1.04) | 0.13 | 0.87 (0.53-1.43) | 0.58 | 1.10 (0.71-1.71) | 0.67 | 0.92 (0.74-1.16) | 0.49 | 2.56 (1.50-4.36) | 0.001 |
| Hepatic failure | 1 | 0.77 (0.58-1.02) | 0.07 | 0.92 (0.43-1.94) | 0.82 | 0.49 (0.18-1.33) | 0.16 | 0.85 (0.59-1.21) | 0.37 | 2.09 (0.86-5.08) | 0.1 |
| **Cancer** | | | | | | | | | | | |
| Liver cancer | 1 | 1.26 (1.01-1.58) | 0.04 | 1.13 (0.60-2.13) | 0.71 | 0.89 (0.44-1.81) | 0.75 | 1.06 (0.78-1.45) | 0.69 | 1.17 (0.37-3.66) | 0.79 |
| **Musculoskeletal** | | | | | | | | | | | |
| Joint replacement surgery (any) | 1 | 0.99 (0.96-1.02) | 0.55 | 0.97 (0.88-1.06) | 0.47 | 1.04 (0.95-1.14) | 0.34 | 0.99 (0.94-1.03) | 0.51 | 1.18 (1.01-1.38) | 0.04 |
| Osteoarthritis | 1 | 0.99 (0.94-1.05) | 0.71 | 1.06 (0.91-1.23) | 0.46 | 1.10 (0.95-1.27) | 0.21 | 1.02 (0.95-1.10) | 0.6 | 1.44 (1.14-1.82) | 0.002 |
| Fractures (any) | 1 | 1.01 (0.97-1.04) | 0.65 | 0.91 (0.82-1.01) | 0.07 | 1.01 (0.92-1.12) | 0.79 | 1.02 (0.97-1.07) | 0.42 | 1.00 (0.84-1.20) | 0.96 |
| Fragility fractures | 1 | 1.02 (0.98-1.07) | 0.38 | 0.87 (0.76-1.00) | 0.05 | 1.10 (0.97-1.24) | 0.13 | 0.99 (0.93-1.05) | 0.74 | 0.85 (0.66-1.09) | 0.19 |
| Osteoporosis | 1 | 0.94 (0.90-0.99) | 0.01 | 0.94 (0.83-1.07) | 0.34 | 0.89 (0.79-1.01) | 0.08 | 0.99 (0.94-1.05) | 0.75 | 1.28 (1.06-1.56) | 0.01 |
| Rheumatoid arthritis | 1 | 0.96 (0.88-1.04) | 0.28 | 0.94 (0.75-1.17) | 0.58 | 1.01 (0.82-1.26) | 0.9 | 0.89 (0.80-0.99) | 0.03 | 1.04 (0.71-1.53) | 0.84 |
| **Brain** | | | | | | | | | | | |
| Any brain outcome (dementia, delirium, or Parkinson's disease) | 1 | 1.02 (0.97-1.09) | 0.43 | 0.93 (0.79-1.11) | 0.43 | 1.03 (0.88-1.21) | 0.72 | 1.00 (0.92-1.08) | 0.9 | 1.30 (1.01-1.68) | 0.04 |
| Dementia | 1 | 1.00 (0.92-1.08) | 0.94 | 0.80 (0.63-1.02) | 0.07 | 1.00 (0.81-1.25) | 0.98 | 1.01 (0.91-1.12) | 0.86 | 1.16 (0.81-1.66) | 0.41 |
| Alzheimer's disease | 1 | 1.08 (0.97-1.20) | 0.18 | 0.87 (0.63-1.20) | 0.4 | 1.22 (0.92-1.61) | 0.17 | 1.02 (0.88-1.17) | 0.84 | 1.04 (0.62-1.77) | 0.88 |

| | | | | | | | | | | |
|---|---|---|---|---|---|---|---|---|---|---|
| Non-Alzheimer's dementia | 1 | 0.93 (0.83-1.04) | 0.2 | 0.72 (0.50-1.03) | 0.07 | 0.81 (0.57-1.13) | 0.21 | 1.03 (0.89-1.18) | 0.73 | 1.27 (0.76-2.05) | 0.33 |
| Delirium | 1 | 1.06 (0.98-1.15) | 0.13 | 1.05 (0.84-1.31) | 0.67 | 1.06 (0.85-1.32) | 0.6 | 1.01 (0.91-1.13) | 0.8 | 1.50 (1.08-2.08) | 0.02 |
| Parkinson's disease | 1 | 0.93 (0.80-1.07) | 0.29 | 0.97 (0.66-1.44) | 0.89 | 1.14 (0.79-1.65) | 0.48 | 0.98 (0.81-1.18) | 0.84 | 1.00 (0.50-2.00) | 0.99 |
| **Pancreas** | | | | | | | | | | | |
| T1 or T2 diabetes | 1 | 1.01 (0.97-1.06) | 0.55 | 0.98 (0.86-1.11) | 0.77 | 1.09 (0.97-1.23) | 0.17 | 1.01 (0.96-1.08) | 0.6 | 1.18 (0.95-1.46) | 0.13 |
| **Infection** | | | | | | | | | | | |
| COVID-19 | 1 | 1.02 (0.94-1.11) | 0.66 | 1.03 (0.81-1.30) | 0.83 | 1.02 (0.80-1.29) | 0.9 | 1.06 (0.94-1.18) | 0.34 | 1.00 (0.65-1.54) | 0.99 |
| Cholecystitis | 1 | 0.98 (0.90-1.08) | 0.73 | 0.92 (0.71-1.18) | 0.52 | 0.99 (0.78-1.26) | 0.92 | 0.99 (0.88-1.11) | 0.86 | 1.49 (1.04-2.13) | 0.03 |
| Pneumonia | 1 | 1.01 (0.96-1.05) | 0.83 | 1.02 (0.90-1.16) | 0.72 | 1.06 (0.93-1.20) | 0.4 | 1.03 (0.97-1.10) | 0.29 | 1.05 (0.83-1.32) | 0.69 |
| Sepsis | 1 | 0.98 (0.94-1.01) | 0.23 | 0.97 (0.88-1.08) | 0.61 | 1.05 (0.95-1.15) | 0.36 | 1.04 (0.99-1.09) | 0.12 | 1.13 (0.96-1.35) | 0.15 |
| LRTI | 1 | 0.95 (0.89-1.01) | 0.08 | 0.92 (0.78-1.09) | 0.34 | 0.93 (0.79-1.10) | 0.39 | 1.04 (0.97-1.12) | 0.3 | 1.16 (0.89-1.52) | 0.28 |
| URTI | 1 | 1.12 (0.98-1.30) | 0.1 | 0.86 (0.56-1.33) | 0.51 | 0.84 (0.54-1.30) | 0.43 | 1.12 (0.93-1.34) | 0.24 | 0.28 (0.07-1.12) | 0.07 |
| UTI | 1 | 0.97 (0.94-1.01) | 0.18 | 0.99 (0.89-1.10) | 0.82 | 1.01 (0.91-1.13) | 0.8 | 1.03 (0.98-1.08) | 0.23 | 1.19 (0.99-1.41) | 0.06 |
| SSTI | 1 | 1.01 (0.95-1.07) | 0.79 | 1.05 (0.90-1.23) | 0.54 | 1.00 (0.85-1.18) | 0.97 | 1.03 (0.95-1.11) | 0.52 | 1.20 (0.92-1.58) | 0.18 |
| **Cardiovascular** | | | | | | | | | | | |
| Arrhythmia | 1 | 1.02 (0.93-1.11) | 0.69 | 0.76 (0.57-1.01) | 0.06 | 1.04 (0.81-1.33) | 0.75 | 0.98 (0.87-1.11) | 0.78 | 1.07 (0.69-1.67) | 0.75 |
| Cardiomyopathy | 1 | 0.97 (0.83-1.13) | 0.71 | 0.82 (0.52-1.30) | 0.41 | 1.11 (0.75-1.64) | 0.61 | 1.08 (0.90-1.31) | 0.41 | 0.44 (0.14-1.37) | 0.16 |
| CHD | 1 | 1.00 (0.95-1.04) | 0.82 | 1.03 (0.92-1.16) | 0.61 | 1.01 (0.90-1.14) | 0.86 | 1.01 (0.96-1.07) | 0.71 | 0.94 (0.76-1.17) | 0.6 |
| Heart failure | 1 | 1.04 (0.97-1.10) | 0.27 | 0.92 (0.77-1.10) | 0.35 | 1.12 (0.96-1.32) | 0.15 | 1.06 (0.98-1.15) | 0.16 | 1.34 (1.03-1.75) | 0.03 |
| **Mental Health** | | | | | | | | | | | |
| Depression | 1 | 0.97 (0.93-1.02) | 0.2 | 1.10 (0.97-1.24) | 0.14 | 1.11 (0.99-1.25) | 0.09 | 1.03 (0.97-1.09) | 0.38 | 0.90 (0.70-1.15) | 0.39 |

HR (Hazard ratio) compared to those with neither *HFE* mutation. Cox proportional hazards regression models adjusted for age, assessment centre, and genetic principal components 1–10. Abbreviations: CHD, coronary heart disease; T1, type 1; T2, type 2; LRTI, lower respiratory tract infection; URTI, upper respiratory tract infection; UTI, urinary tract infection SSTI, skin and soft tissue infection; CI, confidence interval. Joint replacement surgery variable includes a diagnosis of hip, knee, ankle, or shoulder replacement. Any brain outcome variable includes a diagnosis of dementia, delirium, or Parkinson's disease.

**eTable 11**. Incident hospital diagnoses in female UK Biobank participants by p.C282Y/H63D genotypes, excluding a diagnosis of hemochromatosis at baseline

| **Females** | **No mutations** | **H63D +/-** | **H63D +/+** | **C282Y +/ H36D +** | **C282Y +/-** | **C282Y +/+** |
|---|---|---|---|---|---|---|
| Hemochromatosis | 44 (9.24) | 35 (7.35) | 12 (2.52) | 57 (11.97) | 37 (7.77) | 291 (61.13) |
| All-cause mortality | 9847 (59.47) | 3810 (23.01) | 363 (2.19) | 390 (2.36) | 2020 (12.20) | 127 (0.77) |
| **Liver** | | | | | | |
| Liver disease (any) | 3821 (58.38) | 1511 (23.09) | 146 (2.23) | 177 (2.70) | 825 (12.61) | 65 (0.99) |
| Alcoholic liver disease | 180 (52.94) | 78 (22.94) | 11 (3.24) | 6 (1.76) | 59 (17.35) | 6 (1.76) |
| Fibrosis & Cirrhosis | 473 (61.03) | 162 (20.90) | 16 (2.06) | 21 (2.71) | 89 (11.48) | 14 (1.81) |
| Hepatic failure | 199 (64.40) | 60 (19.42) | 7 (2.27) | 4 (1.29) | 35 (11.33) | 4 (1.29) |
| **Cancer** | | | | | | |
| Liver cancer | 232 (55.77) | 114 (27.40) | 10 (2.40) | 8 (1.92) | 49 (11.78) | 3 (0.72) |
| **Musculoskeletal** | | | | | | |
| Joint replacement surgery (any) | 12355 (59.84) | 4771 (23.11) | 454 (2.20) | 500 (2.42) | 2415 (11.70) | 150 (0.73) |
| Osteoarthritis | 4343 (59.08) | 1681 (22.87) | 177 (2.41) | 189 (2.57) | 893 (12.15) | 68 (0.93) |
| Fractures (any) | 10665 (59.31) | 4204 (23.38) | 375 (2.09) | 430 (2.39) | 2188 (12.17) | 119 (0.66) |
| Fragility fractures | 6308 (59.29) | 2517 (23.66) | 214 (2.01) | 275 (2.58) | 1264 (11.88) | 61 (0.57) |
| Osteoporosis | 6990 (60.50) | 2580 (22.33) | 254 (2.20) | 244 (2.11) | 1389 (12.02) | 97 (0.84) |
| Rheumatoid arthritis | 2188 (61.05) | 818 (22.82) | 79 (2.20) | 86 (2.40) | 390 (10.88) | 23 (0.64) |
| **Brain** | | | | | | |
| Any brain outcome (dementia, delirium, or Parkinson's disease) | 3875 (59.16) | 1547 (23.62) | 142 (2.17) | 154 (2.35) | 777 (11.86) | 55 (0.84) |
| Dementia | 2159 (59.72) | 837 (23.15) | 68 (1.88) | 83 (2.30) | 440 (12.17) | 28(0.77) |
| Alzheimer's disease | 1072 (58.20) | 450 (24.43) | 37 (2.01) | 50 (2.71) | 220 (11.94) | 13 (0.71) |
| Non-Alzheimer's dementia | 1099 (61.02) | 395 (21.93) | 31 (1.72) | 34(1.89) | 227 (12.60) | 15 (0.83) |
| Delirium | 1982 (58.17) | 823 (24.16) | 81 (2.38) | 82 (2.41) | 406 (11.92) | 33 (0.97) |

| | | | | | | |
|---|---|---|---|---|---|---|
| Parkinson’s disease | 691 (60.72) | 249 (21.88) | 26 (2.28) | 30 (2.64) | 134 (11.78) | 8 (0.70) |
| **Pancreas** | | | | | | |
| T2 diabetes | 6538 (59.27) | 2575 (23.35) | 240 (2.18) | 275 (2.49) | 1322 (11.99) | 80 (0.73) |
| **Infection** | | | | | | |
| Covid-19 | 1809 (58.83) | 719 (23.38) | 72 (2.34) | 72 (2.34) | 383 (12.46) | 20 (0.65) |
| Cholecystitis | 1749 (59.61) | 675 (23.01) | 62 (2.11) | 68 (2.32) | 351 (11.96) | 29 (0.99) |
| Pneumonia | 6127 (59.06) | 2406 (23.19) | 242 (2.33) | 254 (2.45) | 1275 (12.29) | 70 (0.67) |
| Sepsis | 10024 (59.37) | 3837 (22.72) | 379 (2.24) | 414 (2.45) | 2103 (12.45) | 128 (0.76) |
| LRTI | 3875 (59.94) | 1437 (22.23) | 139 (2.15) | 143 (2.21) | 821 (12.70) | 50 (0.77) |
| URTI | 636 (57.87) | 277 (25.20) | 21 (1.91) | 21 (1.91) | 142 (12.92) | 2 (0.18) |
| UTI | 9137 (59.45) | 3490 (22.71) | 351 (2.28) | 366 (2.38) | 1905 (12.40) | 119 (0.77) |
| SSTI | 3900 (59.15) | 1533 (23.25) | 158 (2.40) | 152 (2.31) | 799 (12.12) | 51 (0.77) |
| **Cardiovascular** | | | | | | |
| Arrhythmia | 1654 (59.75) | 656 (23.70) | 48 (1.73) | 67 (2.42) | 323 (11.67) | 20 (0.72) |
| Cardiomyopathy | 608 (59.90) | 230 (22.66) | 19 (1.87) | 25 (2.46) | 130 (12.81) | 3 (0.30) |
| CHD | 7401 (59.49) | 2875 (23.11) | 294 (2.36) | 291 (2.34) | 1502 (12.07) | 77 (0.62) |
| Heart failure | 3550 (58.50) | 1430 (23.57) | 127 (2.09) | 154 (2.54) | 753 (12.41) | 54 (0.89) |
| **Mental Health** | | | | | | |
| Depression | 6414 (59.58) | 2430 (22.57) | 269 (2.50) | 278 (2.58) | 1317 (12.23) | 57 (0.53) |

Incident disease numbers exclude prevalent disease at baseline. Numbers presented are n (%). Abbreviations: CHD, coronary heart disease; T1, type 1; T2, type 2; LRTI, lower respiratory tract infection; URTI, upper respiratory tract infection; UTI, urinary tract infection; SSTI, skin and soft tissue infection. Joint replacement surgery variable includes a diagnosis of hip, knee, ankle, or shoulder replacement. Any brain outcome variable includes a diagnosis of dementia, delirium, or Parkinson’s disease.

**eTable 12.** Hazard ratios of incident disease outcomes in p.C282Y/H63D genotypes in females, excluding a diagnosis of hemochromatosis at baseline

| **Females** | **No mutations (Ref group)** | **H63D +/-** | | **H63D +/+** | | **C282Y +/ H36D +** | | **C282Y +/-** | | **C282Y +/+** | |
|---|---|---|---|---|---|---|---|---|---|---|---|
| | | HR (95% CI) | P | HR (95% CI) | P | HR (95% CI) | P | HR (95% CI) | P | HR (95% CI) | P |
| Hemochromatosis | 1 | 2.04 (1.31-3.17) | **1.70E-03** | 7.10 (3.75-13.45) | **1.80E-09** | 32.79 (22.10-48.64) | **2.10E-67** | 4.15 (2.68-6.44) | **1.80E-10** | 675.18 (489.89-930.55) | **0.00E+00** |
| All-cause mortality | 1 | 0.99 (0.96-1.03) | 0.69 | 0.95 (0.85-1.05) | 0.31 | 1.00 (0.90-1.11) | 0.98 | 1.01 (0.97-1.06) | 0.61 | 1.13 (0.95-1.35) | 0.16 |
| **Liver** | | | | | | | | | | | |
| Liver disease (any) | 1 | 1.01 (0.95-1.07) | 0.73 | 1.00 (0.84-1.18) | 0.97 | 1.18 (1.01-1.37) | **0.03** | 1.08 (1.00-1.16) | 0.06 | 1.58 (1.23-2.02) | **2.80E-04** |
| Alcoholic liver disease | 1 | 1.10 (0.84-1.43) | 0.48 | 1.57 (0.85-2.88) | 0.15 | 0.79 (0.35-1.78) | 0.57 | 1.54 (1.14-2.06) | **0.43** | 2.75 (1.21-6.20) | **0.02** |
| Fibrosis & Cirrhosis | 1 | 0.87 (0.73-1.04) | 0.13 | 0.87 (0.53-1.44) | 0.60 | 1.11 (0.72-1.72) | 0.65 | 0.93 (0.74-1.16) | 0.51 | 2.67 (1.57-4.55) | **3.00E-04** |
| Hepatic failure | 1 | 0.77 (0.58-1.03) | 0.08 | 0.92 (0.43-1.95) | 0.83 | 0.50 (0.18-1.34) | 0.17 | 0.85 (0.59-1.22) | 0.38 | 1.76 (0.65-4.74) | 0.26 |
| **Cancer** | | | | | | | | | | | |
| Liver cancer | 1 | 1.26 (1.10-1.58) | **0.04** | 1.13 (0.60-2.13) | 0.71 | 0.89 (0.44-1.81) | 0.75 | 1.06 (0.78-1.45) | 0.69 | 1.22 (0.39-3.81) | 0.73 |
| **Musculoskeletal** | | | | | | | | | | | |
| Joint replacement surgery (any) | 1 | 0.99 (0.96-1.02) | 0.54 | 0.97 (0.88-1.06) | 0.47 | 1.04 (0.95-1.14) | 0.35 | 0.99 (0.94-1.03) | 0.51 | 1.16 (0.99-1.37) | **0.07** |
| Osteoarthritis | 1 | 0.99 (0.94-1.05) | 0.72 | 1.06 (0.91-1.23) | 0.46 | 1.09 (0.95-1.27) | 0.22 | 1.02 (0.95-1.10) | 0.59 | 1.46 (1.15-1.85) | **2.10E-03** |
| Fractures (any) | 1 | 1.01 (0.97-1.05) | 0.65 | 0.91 (0.82-1.01) | 0.07 | 1.02 (0.92-1.12) | 0.76 | 1.02 (0.97-1.07) | 0.42 | 1.01 (0.84-1.20) | 0.95 |
| Fragility fractures | 1 | 1.02 (0.98-1.07) | 0.37 | 0.87 (0.76-1.00) | **0.05** | 1.10 (0.98-1.24) | 0.12 | 0.99 (0.93-1.05) | 0.73 | 0.86 (0.67-1.10) | 0.23 |
| Osteoporosis | 1 | 0.94 (0.90-0.99) | **0.01** | 0.94 (0.83-1.07) | 0.34 | 0.89 (0.78-1.01) | 0.08 | 0.99 (0.93-1.05) | 0.73 | 1.26 (1.03-1.54) | **0.02** |
| Rheumatoid arthritis | 1 | 0.96 (0.88-1.04) | 0.28 | 0.94 (0.75-1.17) | 0.58 | 1.00 (0.81-1.25) | 0.97 | 0.89 (0.80-0.99) | **0.03** | 0.96 (0.63-1.44) | 0.83 |
| **Brain** | | | | | | | | | | | |
| Any brain outcome (dementia, delirium, or Parkinson's disease) | 1 | 1.02 (0.97-1.09) | 0.43 | 0.93 (0.79-1.11) | 0.43 | 1.03 (0.87-1.20) | 0.76 | 1.00 (0.92-1.08) | 0.91 | 1.23 (0.94-1.61) | 0.13 |
| Dementia | 1 | 1.00 (0.92-1.08) | 0.94 | 0.80 (0.63-1.02) | 0.07 | 0.99 (0.80-1.24) | 0.95 | 1.01 (0.91- 1.12) | 0.86 | 1.10 (0.76-1.60) | 0.61 |
| Alzheimer's disease | 1 | 1.08 (0.97-1.20) | 0.18 | 0.86 (0.63-1.21) | 0.40 | 1.20 (0.90-1.59) | 0.21 | 1.02 (0.88-1.17) | 0.84 | 1.02 (0.59-1.76) | 0.95 |

| | | | | | | | | | | | |
|---|---|---|---|---|---|---|---|---|---|---|---|
| Non-Alzheimer's dementia | 1 | 0.93 (0.83-1.04) | 0.20 | 0.72 (0.50-1.03) | 0.07 | 0.81 (0.57-1.14) | 0.22 | 1.03 (0.89-1.18) | 0.73 | 1.17 (0.71-1.96) | 0.54 |
| Delirium | 1 | 1.07 (0.98-1.16) | 0.13 | 1.05 (0.84-1.31) | 0.67 | 1.06 (0.85-1.33) | 0.58 | 1.01 (0.91-1.13) | 0.79 | 1.44 (1.02-2.03) | **0.04** |
| Parkinson's disease | 1 | 0.93 (0.80-1.07) | 0.29 | 0.97 (0.66-1.44) | 0.89 | 1.15 (0.79-1.65) | 0.47 | 0.98 (0.81-1.18) | 0.84 | 1.04 (0.52-2.10) | 0.91 |
| **Pancreas** | | | | | | | | | | | |
| T1 or T2 diabetes | 1 | 1.01 (0.97-1.06) | 0.54 | 0.98 (0.86-1.11) | 0.72 | 1.08 (0.96-1.22) | 0.20 | 1.02 (0.96-1.08) | 0.59 | 1.13 (0.90-1.41) | 0.28 |
| **Infection** | | | | | | | | | | | |
| COVID-19 | 1 | 1.02 (0.93-1.11) | 0.68 | 1.03 (0.81-1.30) | 0.83 | 1.02 (0.80-1.29) | 0.88 | 1.06 (0.94-1.18) | 0.34 | 1.00 (0.64-1.56) | 1.00 |
| Cholecystitis | 1 | 0.98 (0.90-1.08) | 0.73 | 0.92 (0.71-1.19) | 0.52 | 0.98 (0.77-1.24) | 0.84 | 0.99 (0.88-1.11) | 0.86 | 1.49 (1.03-2.16) | **0.03** |
| Pneumonia | 1 | 1.00 (0.96-1.05) | 0.84 | 1.02 (0.90-1.17) | 0.71 | 1.05 (0.93-1.20) | 0.41 | 1.03 (0.97-1.10) | 0.29 | 1.02 (0.81-1.29) | 0.85 |
| Sepsis | 1 | 0.98 (0.94-1.01) | 0.22 | 0.97 (0.88-1.08) | 0.62 | 1.05 (0.95-1.16) | 0.34 | 1.04 (0.99-1.09) | 0.11 | 1.14 (0.96-1.36) | 0.14 |
| LRTI | 1 | 0.95 (0.89-1.01) | 0.07 | 0.92 (0.78-1.09) | 0.34 | 0.93 (0.78-1.09) | 0.36 | 1.04 (0.97-1.12) | 0.29 | 1.14 (0.86-1.51) | 0.35 |
| URTI | 1 | 1.12 (0.97-1.29) | 0.13 | 0.86 (0.56-1.33) | 0.51 | 0.84 (0.54-1.30) | 0.43 | 1.12 (0.93-1.34) | 0.24 | 0.29 (0.07-1.17) | 0.08 |
| UTI | 1 | 0.97 (0.94-1.01) | 0.17 | 0.99 (0.89-1.10) | 0.82 | 1.02 (0.92-1.13) | 0.76 | 1.03 (0.98-1.08) | 0.22 | 1.17 (0.97-1.40) | 0.10 |
| SSTI | 1 | 1.01 (0.95-1.07) | 0.81 | 1.05 (0.90-1.23) | 0.54 | 1.00 (0.85-1.17) | 0.98 | 1.03 (0.95-1.11) | 0.52 | 1.20 (0.91-1.59) | 0.19 |
| **Cardiovascular** | | | | | | | | | | | |
| Arrhythmia | 1 | 1.02 (0.93-1.12) | 0.69 | 0.76 (0.57-1.01) | 0.06 | 1.04 (0.82-1.33) | 0.73 | 0.98 (0.87-1.11) | 0.78 | 1.13 (0.72-1.75) | 0.60 |
| Cardiomyopathy | 1 | 0.97 (0.83-1.13) | 0.71 | 0.82 (0.52-1.30) | 0.41 | 1.07 (0.72-1.59) | 0.75 | 1.08 (0.90-1.31) | 0.40 | 0.46 (0.15-1.43) | 0.18 |
| CHD | 1 | 0.99 (0.95-1.04) | 0.81 | 1.03 (0.92-1.16) | 0.60 | 1.01 (0.89-1.13) | 0.92 | 1.01 (0.96-1.07) | 0.70 | 0.92 (0.74-1.15) | 0.48 |
| Heart failure | 1 | 1.03 (0.97-1.10) | 0.28 | 0.92 (0.77-1.10) | 0.36 | 1.12 (0.95-1.32) | 0.17 | 1.06 (0.98-1.15) | 0.15 | 1.35 (1.03-1.77) | **0.03** |
| **Mental Health** | | | | | | | | | | | |
| Depression | 1 | 0.97 (0.93-1.02) | 0.20 | 1.10 (0.97-1.23) | 0.14 | 1.11 (0.98-1.25) | 0.09 | 1.03 (0.97-1.09) | 0.37 | 0.83 (0.64-1.08) | 0.16 |

HR (Hazard ratio) compared to those with neither *HFE* mutation. Cox proportional hazards regression models adjusted for age, assessment centre, and genetic principal components 1–10. Abbreviations: CHD, coronary heart disease; T1, type 1; T2, type 2; LRTI, lower respiratory tract infection; URTI, upper respiratory tract infection; UTI, urinary tract infection SSTI, skin and soft tissue infection; CI, confidence interval. Joint replacement surgery variable includes a diagnosis of hip, knee, ankle, or shoulder replacement. Any brain outcome variable includes a diagnosis of dementia, delirium, or Parkinson's disease

**eFigure 1.** Kaplan-Meier curves for the cumulative incidence of (a) mortality, (b) liver disease, (c) any joint replacement, and (d) any brain outcome, in male *HFE* p.C282Y homozygotes compared to those with no mutations, excluding a diagnosis of hemochromatosis at baseline

a)

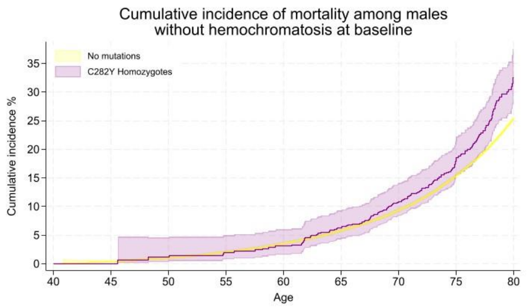


b)

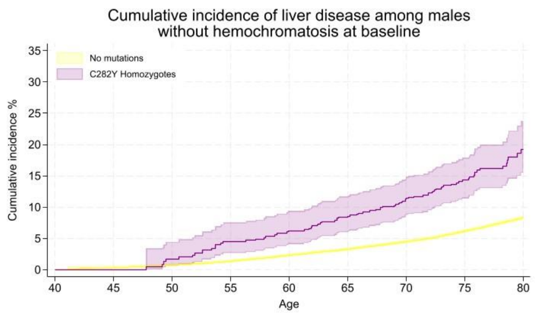


c)

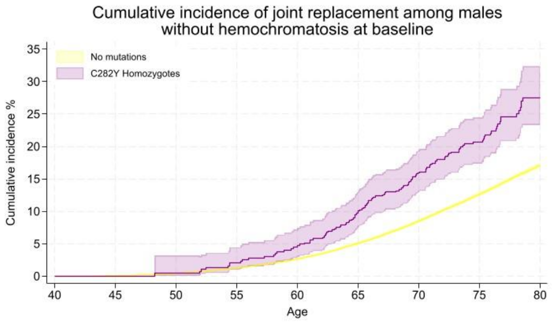


d)

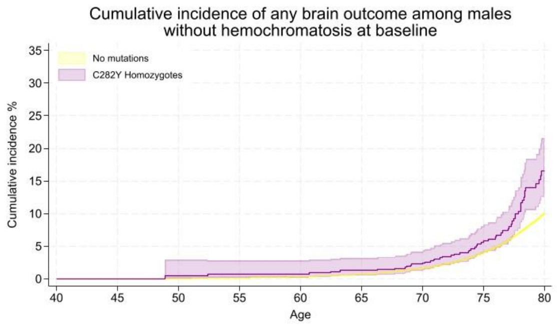


Cumulative incidence (estimated % diagnosed by age 80 years; 95% CIs). Joint replacement surgery includes a diagnosis of hip, knee, ankle, or shoulder replacement. Any brain outcome included a diagnosis of delirium, dementia, or Parkinson’s disease.

**eFigure 2.** Kaplan-Meier curves for the cumulative incidence of (a) liver disease, and (b) any joint replacement, in female *HFE* p.C282Y homozygotes compared to those with no mutations, excluding a diagnosis of hemochromatosis at baseline

a)

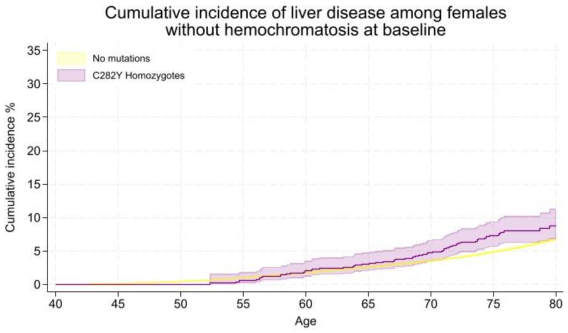


b)

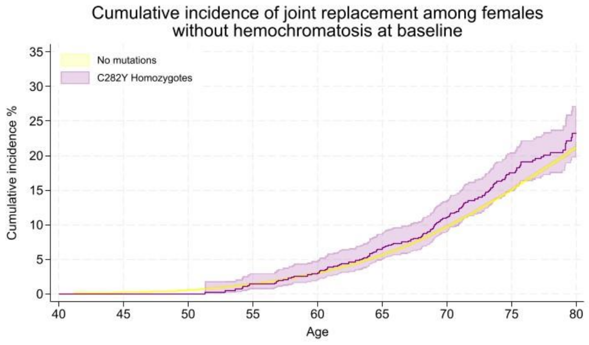


Cumulative incidence (estimated % diagnosed by age 80 years; 95% CIs). Joint replacement surgery includes a diagnosis of hip, knee, ankle, or shoulder replacement.
